# Proteomic and clinical impact of human knockouts in British South Asians

**DOI:** 10.1101/2025.10.15.25337977

**Authors:** Julia Carrasco Zanini, Maik Pietzner, Mine Koprulu, Martijn Zoodsma, Alice Williamson, Karen A. Hunt, Angelos Manolias, Klaudia Walter, Genes & Health Industry Consortium, Genes & Health Research Team, Hilary C. Martin, Sarah Finer, David A van Heel, Claudia Langenberg

## Abstract

Human loss-of-function (LoF) variants affecting both copies of a gene (“human knockouts”) provide a unique opportunity to directly study function and clinical impact of genes but are very rare in most populations sequenced to date. Here we study 1,569 British Bangladeshi and -Pakistani adults who were recalled for plasma sampling for proteomic profiling using three distinct technologies (covering >12,000 proteins) from 55k whole exome sequenced Genes & Health adults – a cohort enriched for rare, biallelic (homozygous) variants due to high autozygosity. We identified 199 individuals with rare homozygous predicted LoF genotypes (pLoF) for which the respective *cis-*protein was measured by at least one technology, and observed extreme (> 3SDs) *cis-*protein underexpression in 41 individuals (median z-score = −9.72 (range: −19.61 to −4.78) and overexpression in 2 individuals (median z-score = 8.1 (range 4.80 – 11.40)), representing 19% of these variants. For missense homozygotes, we observed 158 individuals with significantly under-expressed *cis-*protein (median z-score = −6.95 (range: −28.16 to −3.95)) and 62 individuals over-expressed, median z-score = 5.65 (range: 4.57 to 25.08)). The majority (62%) of LoF knockout genes with an identified cis-protein effect had evidence from 2 or more platforms, highlighting the high confidence nature of these discoveries. Systematic clinical assessment of human knockouts with strong evidence of an impact on *cis-*protein abundance through multi-source electronic health record linkage enabled identification of 1) knockout carriers with rare disease features based on phenotypic similarity, 2) novel rare disease-causing variants, 3) evidence for reclassification of genes and variants of uncertain significance from ClinVar and rare disease panels, and 4) novel gene-phenotype associations in humans. Based on high confidence examples, we developed a machine learning model that predicted 1 in 4 pLOF and 9 in 10 missense variants are likely benign. In summary, our study provides strong human-derived insights into the fundamental biology and clinical relevance of many genes and shows the value of proteogenomic studies of human knockout carriers.

## Introduction

Understanding the function of all genes encoded by the human genome remains one of the main questions in biomedical research. Even over 20 years since the publication of the first reference human genome, the function in humans remains unkown in up to 30% of genes^1^. Disruption of both copies of a given gene in humans, typically through loss-of-function (LoF) variants, but also other deleterious variant classes (e.g. missense variants), enables direct assessment of gene function by studying the phenotypic consequences in humans carrying such variants causing gene “knockouts”^2–5^. However, these deleterious variants are extremely rare in a homozygous state in most populations sequenced so far, and therefore, most of our knowledge on such “human knockout” phenotypes comes from Mendelian recessive disorders. Historically, this has meant individuals with an extreme phenotypic and clinical presentation are the starting point to identify genetic variants responsible for causing rare disorders, i.e. a “top-down” approach (going from phenotype-to-genotype). Sequencing in large-scale population-based studies with linkage to electronic health records (EHRs), only now enables a “bottom-up” approach, where identification of homozygous predicted deleterious variants enables characterisation of the clinical consequences. Such a bottom-up approach can enable systematic characterisation of the entire phenotypic spectrum as opposed to only the most severe end, usually captured by top-down approaches Given the large phenotypic heterogeneity of individual rare disorders, this approach is clinically relevant as it can enable identification of people with often atypical or mixed symptoms caused by poorly understood mutations. However, this is only possible in populations with a high degree of autozygosity that are enriched in rare homozygous predicted LoF variants, commonly resulting from close parental relatedness^3,6^.

Identification of deleterious variants relies on a variety of variant pathogenicity prediction tools which often yield a substantial number of false positives and have limited concordance between them^7^. This is true for classical LoF prediction (e.g. the VEP LOFTEE tool) but especially so for missense variant effect prediction. This uncertainty is amplified for under-represented ethnicities, in part due to the lack of accurate ancestry-specific allele frequency estimates. This results in a large proportion of rare variants (e.g. 36% of ClinVar variants^8^) for which we are unable to judge the molecular and clinical pathogenicity (i.e. variants of uncertain significance or VUS). Complementary functional evidence can resolve VUSs, which can be obtained through animal or cellular models, or RNA sequencing of patient tissue samples. However, current approaches scale poorly (e.g. testing variant impact on cellular models), require invasive tissue biopsies (e.g. bulk tissue transcriptomics from muscle or skin biopsies^9–11^) and may not be useful for variants that only have an impact on the protein product but not the RNA (e.g. missense variants). Broad-capture proteomic technologies now enable simultaneous measurements of thousands of proteins from a single blood sample. Over 80% of proteins that are reliably detected in the circulation are not predicted to be secreted^12^ but can be linked to natural genetic variation (so-called protein quantitative trait loci) acting in cells and eventually affecting their abundance in plasma^13,14^. Therefore, the plasma proteome provides a readout from multiple tissues, that can be used to systematically study the impact of rare variants on the abundance of gene products.

Here, we systematically explore the proteomic and clinical impact of “human knockouts” in an underrepresented population of British Bangladeshi and Pakistani individuals with high rates of consanguinity and hence autozygozity, by integrating data from whole exome sequencing (WES), EHRs and plasma proteomics from three orthogonal platforms targeting over ∼12k proteins.

## Results

We carried out proteomic profiling in plasma samples from 1569 participants from the Genes & Health study^15^ invited back for blood sampling. Participants of this recall study were of similar age (mean 39.95 vs 40.77) and equally likely to be female (55.74% vs 55.49%) but had a higher fraction of the genome with runs of homozygosity (FROH) (0.024 vs 0.018) and were more likely to be Bangladeshi (72.32% vs 57.54), compared to the overall G&H cohort (**Supplementary Table 1**). We performed plasma proteomic measurements using three distinct platforms, including the SomaScan 11K v5 (targeting 9664 unique proteins by 11083 aptamers), the Olink Explore HT (targeting 5416 unique proteins by 5420 assays) and the mass-spectrometry (MS) based Seer Proteograph product suite (5796 unique proteins captured by 5961 protein groups detected in at least 70% of samples). Of 355 (299 high-confidence; 82 low-confidence) and 1363 individuals carrying at least one rare homozygous putative loss-of-function (pLoF: frameshift, stop lost or splice site) or missense variant (minor allele frequency [MAF] < 0.01), i.e. potential human knockouts, we identified 199 (226 variants in 210 genes) and 1246 carriers (7273 variants in 4125 genes) (**see Methods**) for which the protein product of the affected gene (*cis-*protein) was covered by at least one of the three proteomic platforms (**Fig1, Supplementary Table 1**).

**Figure 1.**
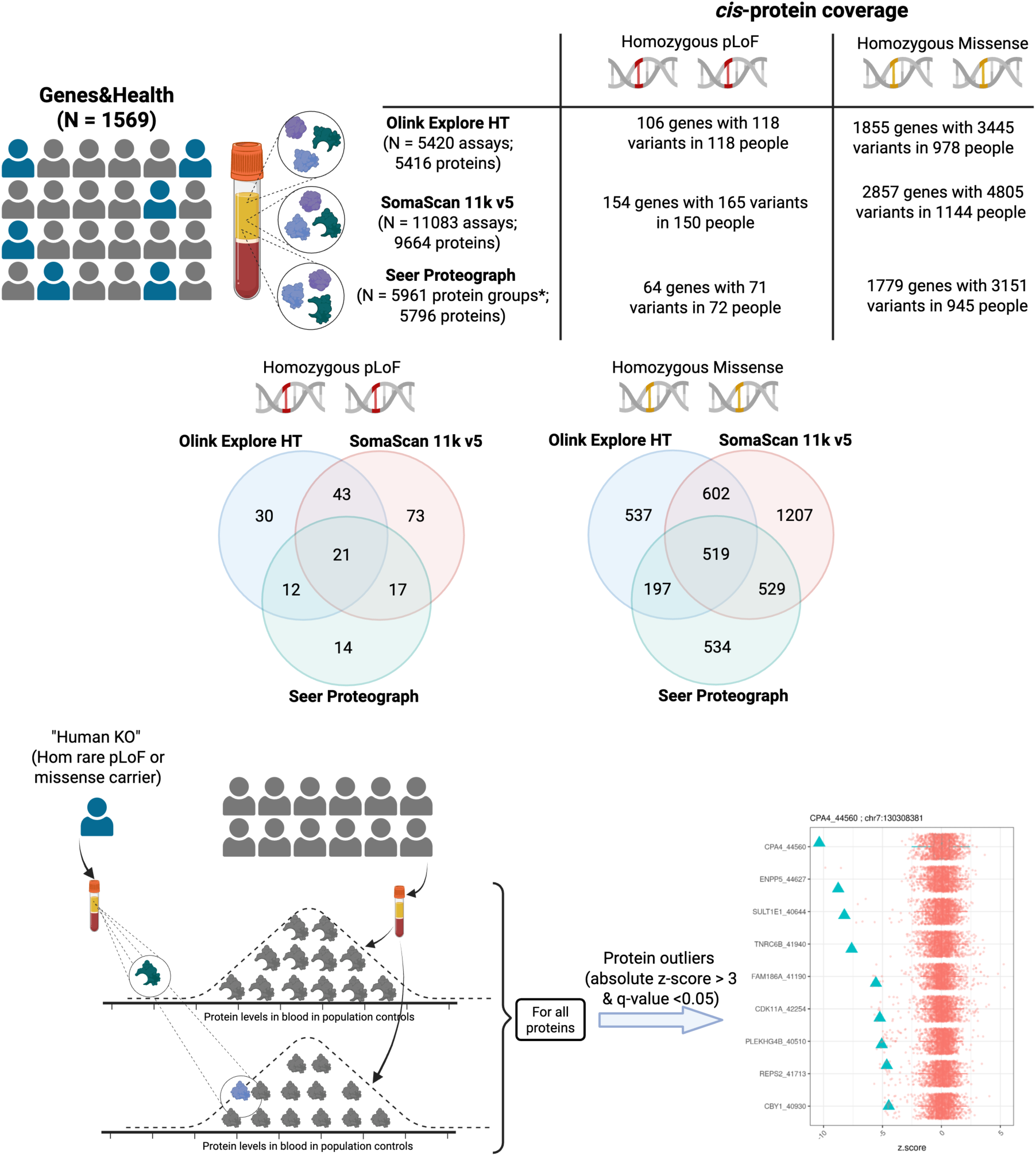
Study design. We performed proteomic profiling in plasma samples from 1569 recalled individuals form the Genes & Health (G&H) study with three different proteomic platforms: Olink Explore HT, SomaScan 11k v5 and Seer Proteograph. We identified homozygous carriers of rare pLoF and missense variants (i.e. potential ‘knockout’ variants) for which the *cis-*protein was covered by at least one proteomic platform. Cross-platform coverage of genes with homozygous pLoF and missense variants is depicted in the in the Venn diagrams. For each homozygous carrier we compared levels of each protein to the rest of the population to identify all protein outliers and whether the *cis-*protein was among these.

### Extreme effects of rare homozygous pLoF and missense variants on *cis-*protein levels

We assessed the effect of rare homozygous pLoF and missense variants on the plasma proteome, by systematically comparing protein abundance in each carrier against all other individuals to identify those significantly (p-value < 0.05/number of proteins in each platform) over- or under-expressed (absolute z-score >3) (**Fig. 1**, **see Methods**). For 42 unique pLoF variants in 43 individuals (19.02% of variants), we observed significant effects on the abundance of the 40 *cis-*protein products (38 under-expressed, median z-score = −9.72 (range: −19.61 to −4.78); 2 over-expressed, median z-score = 8.1 (range 4.80 – 11.40)) (**Fig.2a, Supplementary Table 2**). A total of 62.2% of these were concordant across 2 or more proteomic platforms, while the reminder was seen with only one of the platforms due to single platform coverage (13.3%) or non-consistent effects (24.4%) between platforms. Similarly, we observed significant effects of 227 unique missense variants in 195 individuals (3.53% of variants), on abundance of the 203 *cis-*protein products (158 under-expressed, median z-score = −6.95 (range: −28.16 to −3.95); 62 over-expressed, median z-score = 5.65 (range: 4.57 to 25.08)) (**Fig.2b and Supplementary Table 3**). 32.4% of these were concordant across 2 or more proteomic platforms, while platform specific findings were mostly due to non-consistent effects between platforms (49.3%) rather than missing coverage (18.3%) (**Supplementary Fig. 1**). Our results were consistent with protein under-expression being the expected functional consequence for almost all pLoF variants, as a result of nonsense mediated mRNA decay. In contrast, deleterious missense variants can lead to both protein under- or overexpression through changes in thermodynamic stability, subcellular localization, turnover and complex formation among others ^16,17,18,19^, as evidence by our results.

**Figure 2.**
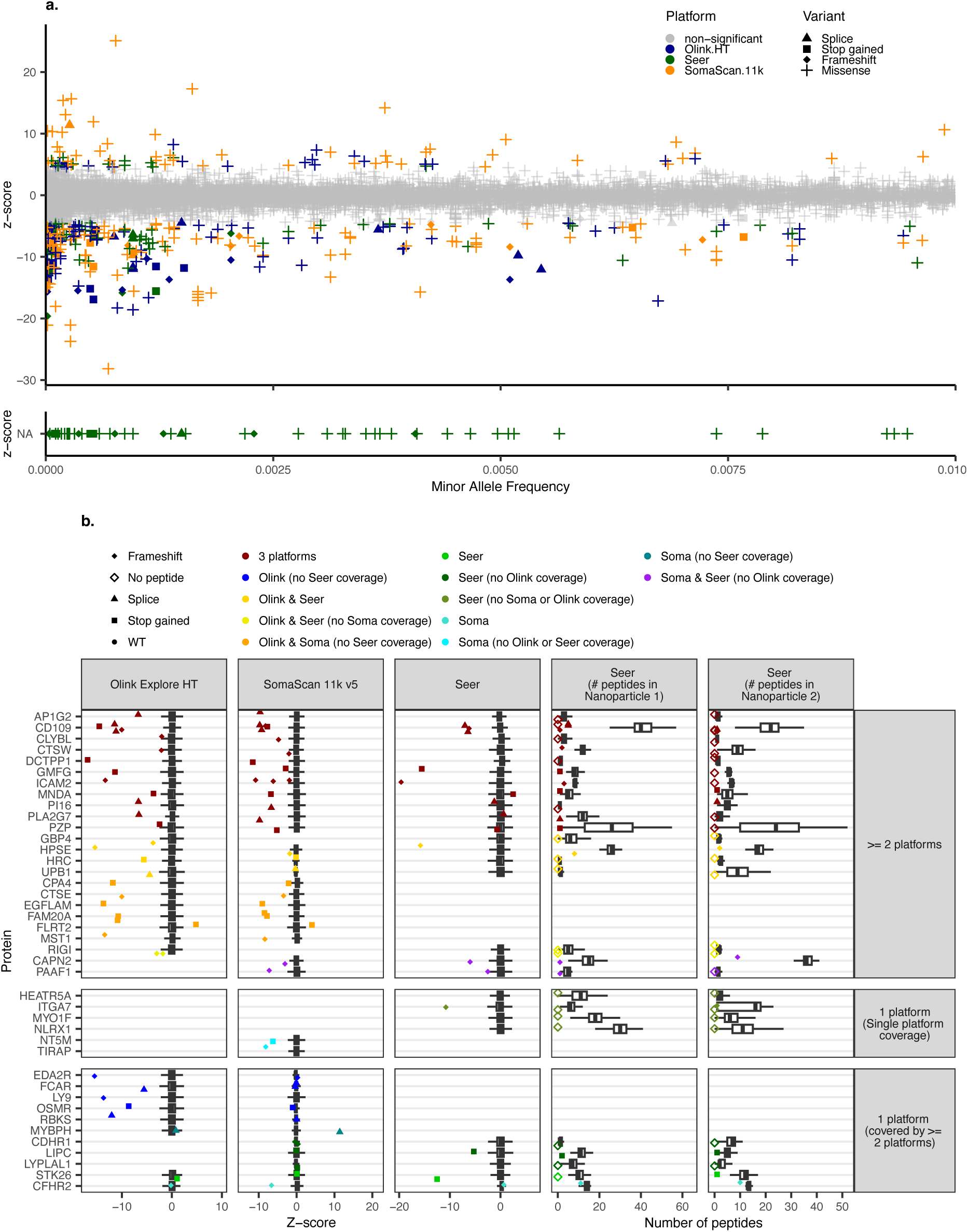
Extreme effects of rare homozygous pLoF and missense variants on *cis-*protein plasma abundance. **a,** Homozygous pLoF and missense variants with evidence of an effect on *cis-*protein abundance across three proteomic platforms (Olink, SomaLogic and Seer). Cis-proteins for which no peptide was detected (i.e. no *cis-*protein detection) in the ‘knockout’ variant carrier but there were at least 5 peptides that were detected in the rest of population are depicted as a z-score of −50. **b,** pLoF variants with evidence of an effect on *cis-*protein abundance detected by at least one proteomic platform. Z-scores represent the number of standard deviations protein levels in the variant carrier are away from the population. For the Seer proteograph platform variant effects were further evaluated on the number of peptides detected in the variant carrier in comparison to the population distribution.

In contrast to affinity-based platforms, the MS-based platform enabled detection of variant effects not only on plasma protein abundance, but also as (likely) absence of detection of the *cis-*protein product in blood (as seen for 11 of 68 variants for which the *cis-*protein was captured by Seer) and through the number of peptides detected for the *cis-*protein product in variant carriers compared to the population (**Fig.2a and Supplementary Fig. 1, see Methods**).

For 51% and 45% of these pLoF and missense variants we also observed evidence of an effect on protein abundance in heterozygous carriers evidenced by an absolute average z-score > 2. We used this less stringent z-score threshold for heterozygous carriers to enable detection of expected smaller effects. Indeed, we found that homozygous carriers of pLoF and missense variants with significant *cis-*protein evidence from at least one platform had an average 6-fold (range: 0.65-fold – 35-fold) greater effect compared to heterozygous carriers (**Supplementary Fig. 2**).

Of 240 genes for which we observed extreme effects of homozygous pLoF or missense variants on *cis-*protein abundance (i.e. protein-informed knockout variants), only ∼30% (n=71) are well established diagnostic genes for rare disorders enabling systematic characterisation of potential clinical consequences in so far poorly characterised genes. Furthermore, even for well-characterised diagnostic genes where the ‘knockout’ variant has a strong impact on the *cis-*protein, absence of the expected disease can result from compensation by genetic canalization. Therefore, we sought to systematically characterise the clinical consequences in protein-informed human knockouts.

### Clinical characterisation of protein-informed homozygous LoF and missense variant carriers

Characterisation of the clinical impact of homozygous LoF and missense variants with evidence of strong *cis-*protein effects (i.e. protein-informed knockout variants), can: 1) enable identification of KO variant carriers with rare disease features, 2) lead to discovery of novel rare disease-causing variants, 3) provide evidence for reclassification of variants and genes of uncertain significance from ClinVar and rare disease panels and 3) lead to discovery of novel gene-phenotype associations in humans.

To gain insights into the clinical consequences of protein-informed knockout variants, we first computed phenotype matching scores^20^, indicating the degree of similarity between established gene-phenotype associations (from sources including OMIM^21^, Orphanet and ClinVar^22^) and phenotypes recorded in electronic health records (EHRs) of the respective carriers (**see Methods**). For only 35% of genes with at least one protein informed human knockout carrier there was prior knowledge on established phenotype links for the affected gene. Of these individuals, more than half (n=79) had a similar clinical presentation to that expected based on prior knowledge (phenotypic matching score ≥ 0.2), determined through manual chart review) for genes with diagnostic-grade (“green”), borderline (“amber”) and low (“red”) evidence of disease causation according to Genomic England’s PanelApp^23^ resource or with mouse knockout evidence only (**Fig3.a**). We observed replication of phenotypic consequences for 74% of these variants in at least one additional carrier from the entire G&H cohort of >55,000 volunteers supporting phenotype matching.

**Figure 3.**
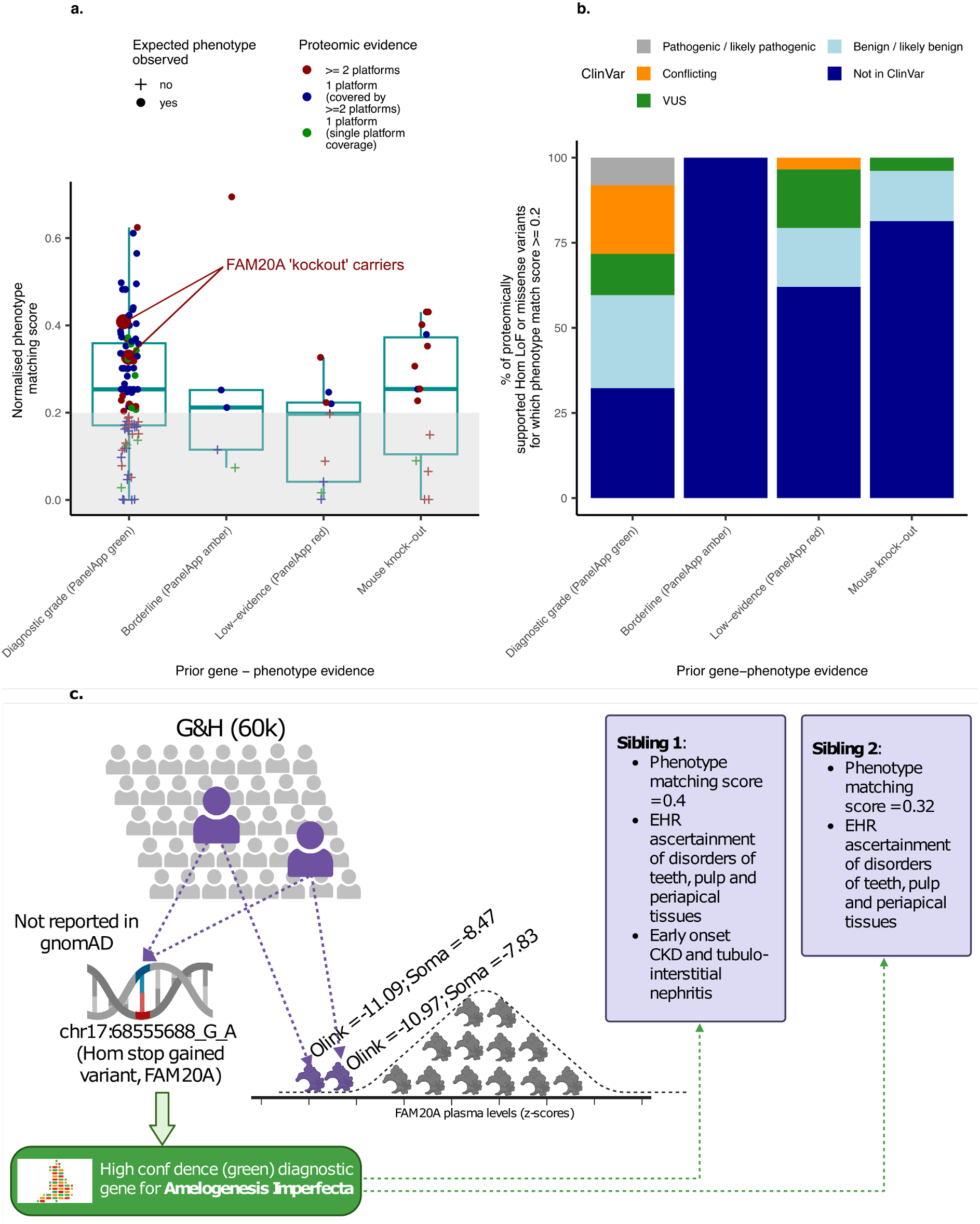
Clinical characterisation of protein-informed human knockouts. **a,** Phenotype matching scores for homozygous LoF or missense variants with evidence of cis-protein under- or over-expression (i.e. protein-informed human knockouts) for which gene-phenotype associations have previously been characterised with diagnostic-grade (PanelApp green genes), borderline (PanelApp amber genes), low (PanelApp red genes) evidence in humans or through mouse knockout models. Results are normalised to the highest phenotype matching score across all individuals, due to the lack of a standard scale. **b,** Summary of ClinVar evidence for protein-informed ‘knockout’ variants in carriers with expected clinical consequences according to previously characterised gene-phenotype associations. **c,** Identification of novel pathogenic variant in the FAM20A gene in siblings with phenotypic evidence matching the clinical presentation of Amelogenesis Imperfecta.

We note that phenotype matching currently ignores the presence of multiple homozygous pLoF variants in the same volunteer that can lead to blended phenotypes ^24^. About a third (n=29) of the participants with converging proteomic and clinical evidence had pLoFs in at least one other gene (mean number of genes with homozygous pLoF variants = 1.73, range: 1 – 6), but only 3 individuals had a higher phenotypic matching score for another “knocked out” gene (independently of whether it was captured by any proteomic platform) (**Supplementary Fig. 3**).

### Identification of novel disease-causing variants and reclassification of pathogenicity of variants in established disease genes

Even in human knockouts for diagnostic-grade genes (i.e. PanelApp green genes) ascertaining whether a clinical diagnosis for a rare disease had been made is challenging, given the lack of specific ICD-10 codes for many of these conditions. A total of 61 homozygous LoF or missense carriers had converging *cis-*protein under-expression and matching clinical characteristics (phenotype matching score ≥ 0.2) for diagnostic-grade genes, including 16 carriers of novel likely pathogenic variants not previously reported in ClinVar ^22^, and therefore likely undiagnosed (median *cis-*protein z-score = −8.03 (minimum = −12.72 to maximum = 13.08)). Among these were also 16, 10 and 15 carriers of variants annotated as benign (median *cis-* protein z-score = −5.19 (−15.71 to 25.07)), VUSs (median *cis-*protein z-score = −5.34 (−10.98 to 10.22)) or with conflicting pathogenicity (median *cis-*protein z-score = −4.81 (−10.76 to 9.85)) annotations in ClinVar (**Fig.3b and Supplementary Table 4**). Among these protein-informed homozygous LoF or missense carriers for diagnostic-grade genes, we identified only 4 carriers of known pathogenic variants reported in ClinVar (median *cis-*protein z-score = −6.86 (−8.00 to −5.22)), of which only 1 had any evidence of having a clinical diagnosis through their EHRs (i.e. an individual with retinal dystrophy with a homozygous frameshift variant, chr10:8421138:AG:A, in *CDHR1*).

Novel pathogenic variants included a homozygous stop gained variant in the *FAM20A* gene (chr17:68555688:G:A) found in two siblings with strong FAM20A protein under-expression detected through the Olink (z-scores = −11.09 and −10.97) and SomaLogic platforms (z-scores = −8.47 and −7.83) (**Fig.3c**). This variant was not present in gnomAD or any other individual of the ∼55,000 with WES in G&H. *FAM20A* is a high confidence causal gene (“green”) for amelogenesis imperfecta, a condition characterised by abnormal enamel formation, a range of dental problems and in rare cases, renal problems resulting in eventual kidney failure^25^. Both siblings (phenotype matching scores = 0.40 and 0.32) had clinical evidence of “diseases of pulp and periapical tissues” (ICD-10 code: K04) and “other disorders of teeth and supporting structures” (ICD-10 code: K08). One of these siblings was additionally diagnosed with chronic kidney disease and tubule-interstitial nephritis in their 20’s.

#### Strengthening gene-phenotype links in humans for genes with insufficient prior evidence

Genes with borderline (PanelApp “amber”) and low (PanelApp “red”) evidence of a disease-causing role are not usually considered for rare disease diagnostics, as additional evidence supporting the gene-phenotype link is needed. Among the 7 protein-informed human knockouts with matching clinical characteristics for borderline and low confidence PanelApp genes, we identified 3 variants (median *cis-*protein z-score = −4.72 (−9.79 to 11.93)) that had been previously reported as VUSs in ClinVar and 4 novel variants not previously reported in ClinVar (**Fig.3b Supplementary Table 4**).

This included a splice donor variant (chr6:46711495:C:T, gnomAD MAF = 4.01×10^-4^) in the *PLA2G7* gene reported as a VUS in ClinVar. This is a low confidence (red in PanelApp) gene for severe multi-system atopic disease with high IgE ^23^ characterised by atopy susceptibility, asthma, recurrent bacterial infections of the skin and eczema^26^. The homozygous carrier for this variant had *cis-*protein under-expression supported by all three platforms (PLA2G7 Olink z-score = −9.77, SomaScan z-score = −9.77, Seer: only 1 peptide detected compared to 11.8 peptides on average in the population) and was diagnosed with asthma and psoriasis (phenotype matching score = 0.22). We identified 7 additional homozygous carriers for the chr6:46711495:C:T *PLA2G7* variant in G&H participants that were not part of this recall study. Among all homozygous carriers, there was modest over-representation of asthma (3 out of the 8 carriers) compared to the background population (fisher exact test odds ratio = 3.55, p-value = 0.09) and phenotypic similarity scores ranged from 0.19 to 0.60 (mean = 0.34) providing further support for a link between *PLA2G7* and atopy susceptibility. Such human genetic evidence may raise concerns about strategies that inhibit PLA2G7 activity to treat cardiovascular disease, or even extend human health span ^27^.

Among variants unreported in ClinVar, the human knockout with the highest phenotypic similarity (phenotype matching score = 0.69) was a homozygous carrier for a frameshift variant (chr3:49687057:GAC:G, gnomAD MAF = 3.8×10^-4^) in the *MST1* gene, supported by Olink (MST1 z-score = −13.69) and SomaLogic (MST1 z-score = −8.39) (no Seer coverage). *MST1* is a medium confidence gene (amber in PanelApp) for Epidermodysplasia verruciformis^23^, a recessively inherited dermatological condition characterised by warty skin lesions and increased risk of malignant skin tumours ^28^. This homozygous pLoF carrier presented with pityriasis-like lesions (commonly seen in Epidermodysplasia verruciformis) and benign overgrowth of the feet’s deep connective tissue (plantar fibroma), accounting for the observed phenotypic similarity. Among the 12 additional homozygous carriers for the chr3:49687057:GAC:G *MST1* variant in the entire G&H cohort (no proteomic data available), phenotype matching scores ranged from 0.06 to 0.70 (mean = 0.40); 8 of which had the expected clinical characteristics (phenotype matching score ≥ 0.2), further supporting the association between *MST1* and Epidermodysplasia verruciformis-like phenotypes.

We further identified a novel homozygous stop gained variant carrier (chr15:58542552:C:T, gnomAD MAF = 3.2×10^-5^, not present in any other G&H individual) in the *LIPC* gene, a medium confidence gene for rare metabolic disorders. Homozygous rare variants in *LIPC* have been described in cases of hepatic lipase deficiency ^29^, a recessive inherited condition characterised by the presence of xanthomas, angina pectoris and premature coronary artery disease in spite of elevated high-density lipoprotein (HDL) and reduced low-density lipoprotein (LDL) levels, given their abnormal composition with a much higher triglyceride content ^30^. The homozygous LoF carrier of this novel variant in *LIPC* had hypertrophic skin lesions (which can resemble xanthomas upon clinical examination^31^). LDL and HDL levels recorded through their EHRs were respectively significantly lower (median z-score across all records = −3.3) and higher compared to the population (median z-score across all records = 2.7), consistent with clinical presentation of hepatic lipase deficiency, but had unremarkable triglyceride levels. Furthermore, metabolomic data from this individual (see methods) showed extremely high levels (z-score > 5) of phosphatidylethanolamine (PE) metabolites (**Supplementary Fig. 4, see Methods**), consistent with LIPC’s function as a phospholipase. The lipid profile in this individual is further consistent with associations identified between rare genetic variation in the LIPC gene and increased triglyceride and phospholipid content in HDL particles through aggregate variant testing in UK Biobank ^32^.

For several protein-informed human knockouts direct gene-phenotype links have only been characterised in mice, while only indirect links are established in humans (i.e. through protein-protein interactions or pathways). Among these, the finding with the highest degree of phenotypic similarity (phenotype matching score = 0.43) was for a homozygous frameshift variant carrier (chr7:73758966-T-TTG, not present in gnomAD or any other G&H individual) in the *CD109* gene. Knockout *CD109* mice display a range of skin, eye and hair phenotypes^33^ as well as asthma susceptibility ^34^ in line with the symptoms present in the human carrier of this ‘knockout’ variant, including urticaria, excessive hair loss and asthma.

### Prioritisation of unreported gene-phenotype links

Among protein-informed human knockouts for which we were unable to generate a phenotype matching score (**see Methods**) we investigated the number of life-time diagnoses using ICD-10 4-digit codes to establish the potential clinical features and health burden. While protein-informed human knockouts had a similar average number of life-time diagnoses compared to individuals with rare homozygous pLoF or missense variants with no effect on the *cis-*protein or those without any knockout variants (Anova p-value = 0.1); we observed examples at both ends of the distribution, i.e. those with no diagnoses and likely no clinical impact^6^, and those with an extremely high number of diagnoses (**Fig.4a**). We identified 7 protein-informed human knockouts without a phenotype matching score among the top 5% of the distribution of lifetime diagnoses (mean = 89, range: 77 to 104 ICD-10 4 digit codes). For 4 of these individuals, we observed concordance between the tissue or organ system most affected (i.e. the ICD-10 chapter with the greatest number of diagnoses) and the cell type with highest expression of the knocked-out gene (**Supplementary Fig. 5 and Supplementary Table 5**). For example, a homozygous splice variant carrier (chr1:203175326:C:A, gnomAD MAF = 1.2×10^-5^, not present in any other G&H individual) in the *MYBPH* gene (SomaScan MYBPH z-score = 11.39) predominantly expressed in skeletal myocytes^35^, had the highest number of musculoskeletal system and connective tissue diagnoses (ICD-10 chapter XIII, **Fig. 4a-b**) compared to other ICD-10 chapters. Aggregate variant testing in UK Biobank and All of Us further showed that heterozygous LoF function variants in *MYPBH* were most strongly associated with disorders of the joints (beta = 0.58, p-value = 1.31×10^-4^) and intervertebral disc disorders (beta = 0.70, p-value = 2.28×10^-4^)^36^, consistent with the range of diagnostic codes reported in the *MYBPH* knockout including spondylosis and arthritis. Paraspinal muscles have a crucial role in spinal stability with studies showing linking paraspinal muscle atrophy with accelerated disc degeneration^37^. While proteomic evidence showed increased MYBPH plasma levels in the human knockout, their clinical characteristics likely indicated this is a true LoF variant, which likely results in increased levels of a dysfunctional protein isoform recognised by the SomaLogic assay. Alternatively, increased plasma levels could indicate defective intracellular binding to Myosin, its native function, leading to spillover into the blood.

**Figure 4.**
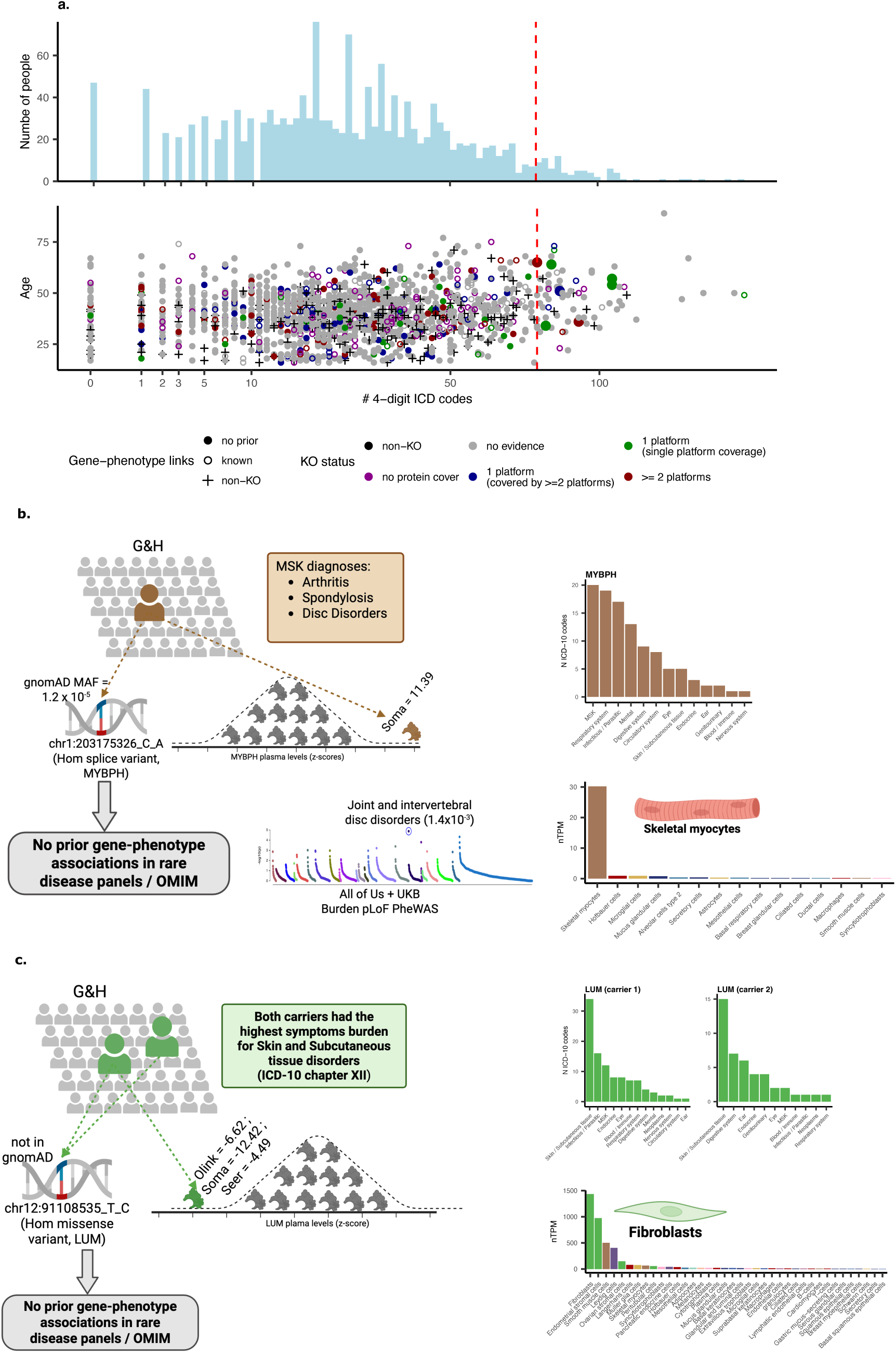
Prioritisation of novel gene-phenotype links in human knockouts. **a,** Distribution of the number of unique diagnoses (4 digit ICD-10 codes) across all individuals with proteomics from the G&H study. Points under the histogram highlight location within the distribution for individuals without any potential ‘knockout’ variants (black), individuals with ‘knockout’ variants with no evidence of an effect on *cis-*protein abundance and protein-informed human knockouts supported by one (blue or green) or multiple platforms (red). Bigger point represents example of individuals with protein-informed knockout variants but no phenotype-matching score in the top 5% of the distribution of number of diagnoses. **b,** Identification of a protein-informed knockout variant in the *MYBPH* gene, in an individual with the greatest symptoms burden for Musculoskeletal (MSK) conditions, consistent with skeletal muscle being the cell type expressing *MYBPH* (data from the Human Protein Atlas ^35^). **c,** Identification of a protein-informed knockout variant in the *LUM* gene, in an individual with the greatest symptoms burden for skin and subcutaneous tissue conditions, consistent with skeletal fibroblasts being the main cell type expressing *LUM* (data from the Human Protein Atlas).

Similarly, a knockout variant carrier in the *LUM* gene that was supported by all proteomic platforms (LUM Olink z-score = −6.62, SomaLogic z-score = −12.41, Seer z-score = −4.49), with a homozygous missense variant in the *LUM* gene (chr12:91108535:T:C, not reported in gnomAD) had the greatest number of diagnostic codes reported for diseases of the skin and subcutaneous tissue (ICD-10 chapter XII). *LUM* has the highest gene expression in fibroblasts ^35^, an abundant cell type in the skin and connective tissues. The only other homozygous chr12:91108535:T:C carrier in the entire G&H cohort also had the greatest number of diagnostic codes for diseases of the skin and subcutaneous tissue (Jaccard index between ICD-10 chapter XII codes between both LUM knockouts = 0.1), further supporting this unspecific gene-phenotype association (**Fig. 4c**). There have further been few reports of an unconfirmed association between *LUM* and severe myopia. Only one of the knockout variant carriers had a relevant diagnosis, for corneal degeneration, in line with the mice knockout phenotype^38^. We observed a similar pattern for two of the other protein-informed knockouts. These male individuals had heterozygous missense variants in the *HEPH* (chrX:66255130:G:A, gnomAD MAF = 8.36×10^-7^) and the *DIPK2B* (chrX:45157746:A:G, gnomAD MAF = 1×10^-5^) genes, which are mostly expressed in enterocytes and endothelial cells, respectively^35^. Consistently, the *HEPH* knockout had the highest number of diagnostic codes for digestive system disorders (ICD-10 chapter XI); while the *DIPK2B* knockout had the highest number of diagnostic codes for circulatory system disorders (ICD-10 chapter IX). One of the 2 additional male heterozygous carriers for the chrX:66255130:G:A variant (*HEPH* knockouts) in the entire G&H cohort, also had the highest number of diagnostic codes for digestive system disorders (Jaccard index between ICD-10 chapter XI codes in both HEPH knockouts = 0.7) (**Supplementary Fig.6a-d**). *HEPH* is a low confidence gene for congenital anaemias and iron metabolism disorders, however, we found no such evidence in the ‘knockout’ carriers (accounting for the lack of mappable HPO terms) but rather a range of unspecific digestive system disorder codes. None of the 2 additional male heterozygous chrX:45157746:A:G variant carriers (*DIPK2B* knockouts) identified in the entire G&H cohort, had the highest symptom burden for circulatory system disorders (**Supplementary Fig.6e-h**). Altogether our results suggest that human knockouts might display a higher unspecific symptoms burden in specific organ systems or tissues which are the main site of expression of the knocked-out gene is most strongly expressed.

### Factors facilitating identification of variants with true extreme effects on *cis-*protein abundance

We demonstrated the value of bottom-up strategies to accelerate characterisation of rare disease gene-phenotype links, but scaling is an inherent limitation. We therefore sought to develop a machine learning model to predict the likelihood of observing an effect of rare homozygous pLoF and missense variants on *cis-*protein plasma abundance. We curated a comprehensive list of 246 variables across 8 different groups of gene, variant, and protein biological and technical characteristics (**Supplementary Table 6, Fig. 5a**). We focused on high-confidence, likely true positive findings with *cis-*protein evidence from at least 2 platforms, which we divided in a 75% training and 25% validation set (**see Methods**). An extreme gradient boosting model (XGBoost) model based on 129 features (**Fig.5a-b**), achieved an area under the receiver operator curve (AUROC) of 0.98 (95% CI 0.96 – 0.99) (**Fig.5c**), with a sensitivity of 0.96 and a specificity of 0.86 at a cutoff of 0.0026. Most of the 129 predictors (52.89%) were nominally associated with an increased likelihood of a variant effect on *cis-*protein abundance (median Odds ratio (OR) = 1.91, range= 1.18 to 45.01), with only 9 features inversely associated (median Odds ratio (OR) = 0.54, range= 0.11 to 0.71). A total of 27 features across 5 groups which were associated with detection of a variant effect on *cis-*protein abundance (FDR < 0.05, **Fig. 5b, Supplementary Table 7**) accounting for factors associated with the efficiency of non-sense mediated RNA decay, such as variant position along coding transcript and exon number and minor allele frequency^39^. Protein assay quality and protein characteristics, contributed the most to the predictive model performance, shown by the superior performance of a model trained on protein characteristics only (i.e. excluding all genetic features) (AUROC = 0.97 (95% CI 0.95 – 0.99)). However, genetic features alone retained strong predictive performance (AUROC = 0.91 (95% CI 0.84 – 0.99)) (**Fig.5c and Supplementary Figure 7**).

**Figure 5.**
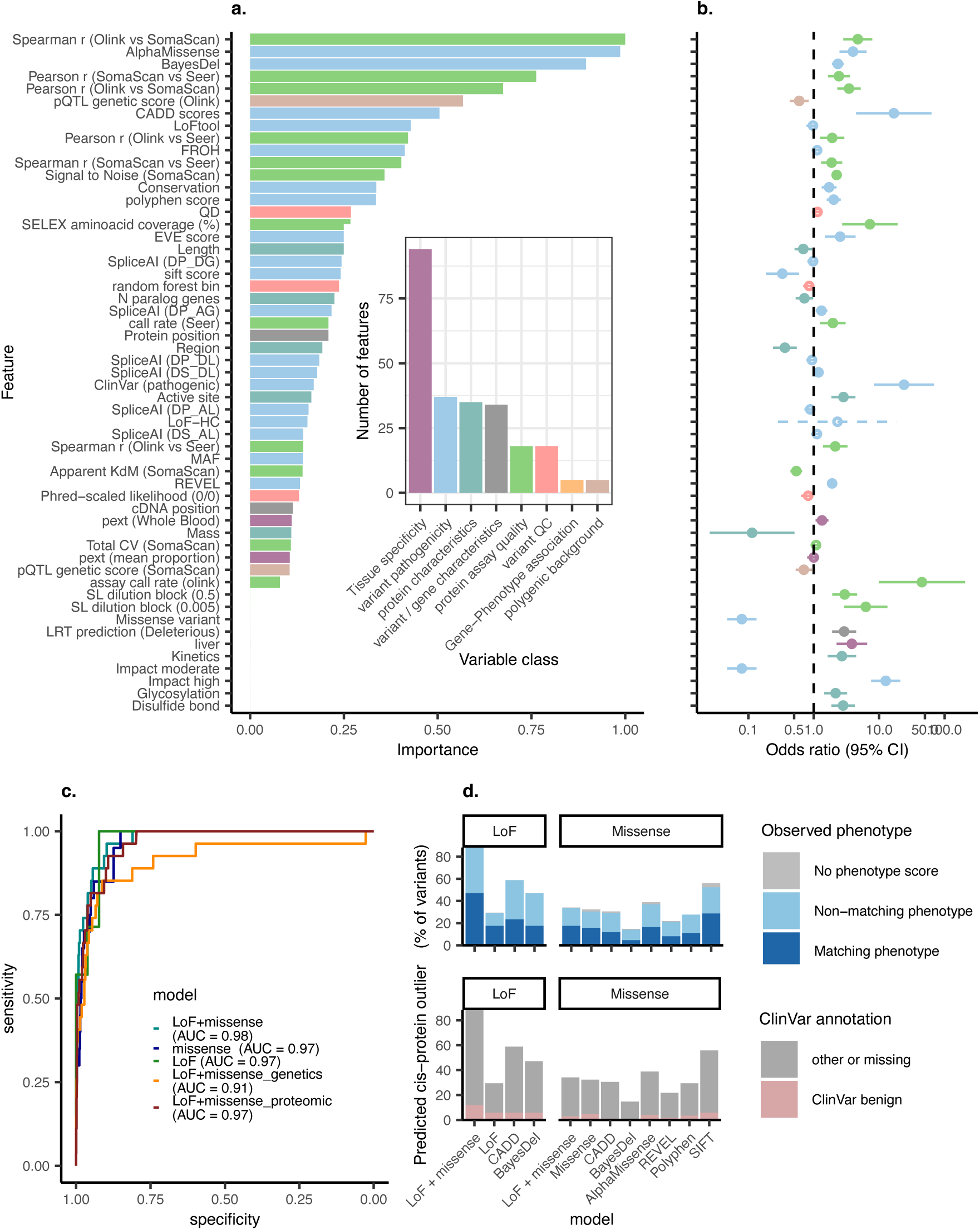
Prediction of protein-informed homozygous LoF and missense human knockouts. **a,** Ranked feature importance (normalised to the feature with the highest score) from an XGBoost model trained to predict detection of *cis-*protein outliers for homozygous LoF and missense variants (LoF + missense model). Inset represents the number of features per group tested. **b,** Associations between features and the likelihood of detecting a cis-protein outlier for all predictive feature or those significantly associated at a 5% FDR threshold. **c,** ROC curves for XGBoost model trained to predict detection of *cis-*protein outliers for homozygous LoF and missense variants (LoF + missense model, including all variables, genetics only or proteomic only variables), LoF only (LoF model) or missense only variants (missense model). **d,** Model testing in homozygous LoF and missense variants with *cis-*protein effects detected through only one platform. Proportion of ‘knockout’ variants predicted to impact *cis-*protein abundance by our developed models or standard variant pathogenicity scores for which a matching phenotype was observed (i.e. likely true positives) or had a benign annotation in ClinVar (i.e. likely false positives).

We similarly trained models to predict the likelihood of *cis-*protein under- or over-expression detection specifically for missense or LoF variants only. These models had a highly comparable performance to the joint LoF+missense model, with the missense only model achieving an AUROC of 0.97 (95% CI 0.94 – 0.99), a sensitivity of 0.95 and a specificity of 0.87 at a cutoff of 0.0015; and the LoF only model achieving an AUROC of 0.97 (95% CI 0.93 – 1), a sensitivity of 1 and a specificity of 0.92 at a cutoff of 0.072 (**Fig.5c**). We identified 22 variables out of 230 gene, variant and protein biological and technical characteristics that were associated with detection a missense variant effect on *cis-*protein abundance (**Supplementary Figure 8c and Supplementary Table 8**). Of those features, only the SpliceAI delta score for donor loss was not identified to be associated with detection of effects on *cis-*protein abundance when considering both LoF and missense variants. None of the variables tested were significantly associated with detection of effects on the *cis-*protein for LoF variants only.

Among informative features included in these models the majority were variant pathogenicity scores, tissue specificity and protein assay quality characteristics (**Fig.5b, Supplementary Figure 9a-b**). Protein assay quality, such as detectability in plasma and cross-platform correlations, were the important predictors. This likely indicates that protein assays that are less robustly measured in plasma or less generalisable across platforms hinder our ability to detect rare protein quantitative loci (pQTLs). As expected, variant pathogenicity scores were also among the strongest predictors. Additional important predictors included features that indicated increasing protein complexity (e.g. length, mass, presence of regions of interest with specified functions or characteristics and active sites among others) as well as additional “modifying” common and rare variants which might rescue or counteract the effect of the homozygous LoF or missense variants on abundance of the *cis-*protein product (e.g. common pQTLs, additional LoF variants in the affected gene). While common pQTLs can also serve as proxies of how well the protein target is measured, we did find evidence of genetic modifying effects (**Supplementary Fig. 10**). Only 17.93%, 16.28% and 2.17% of informative features from the LoF+missense, missense only and LoF only models were specific to the corresponding model.

We applied the optimised models to variants for which we only observed evidence of an effect on *cis-*protein abundance with 1 proteomic platform. Based on the LoF+missense and LoF only prediction model scores, 88.2% and 29.4% of platform-specific *cis-*protein outliers for LoF variants were predicted to be true positives, whereas 34.1% and 32.4% of platform specific *cis-*protein outliers for missense variants were predicted to be true positives by the LoF+missense and missense only models (**Fig.5d**). Given the lack of a ground truth for this set of platform specific findings, we integrated evidence from phenotypic matching scores to assess model sensitivity, as *cis-*protein outliers for LoF and missense variants in people with the expected phenotypic consequences (phenotype matching score ≥ 0.2) are much more likely to represent true positives. However, the absence of phenotypic similarly (phenotype matching score < 0.2) does not translate to model specificity (i.e. how well the model can identify true negatives) given the potential for compensatory mechanisms which can ‘rescue’ the phenotype despite a true functional effect of pathogenic variants on *cis-*protein abundance. Therefore, we tested model specificity through ClinVar benign annotations, which while imperfect provides a reference in the absence of a truth standard, among variants predicted to have a functional effect on *cis-*protein abundance. The LoF+missense XGBoost model was more sensitive compared to most variant pathogenicity scores tested (**see Methods**), except the SIFT score which predicted a larger number of pathogenic variants in individuals with matching phenotypic evidence (**Fig5.d**), albeit at a loss of specificity shown by the greater number benign ClinVar variants among those predicted as pathogenic. The XGBoost LoF+missense model had similar sensitivity to the AlphaMissense score but was more specific. We therefore demonstrate the potential of proteomic data to improve variant pathogenicity prediction. Overall, our XGBoost model had the best performance among all scores tested balancing sensitivity and specificity for missense variants. However, while the sensitivity for LoF variants outperformed all other scores, it was the least specific compared to other scores (**Fig5.d**).

We finally hypothesized that assay sensitivity to detect low abundant proteins in plasma is a contributing factor to the observed low detection rate of *cis-*protein effects for rare homozygous pLoF (∼19%) and missense (∼3%) variants. This was supported by indicators of plasma protein measurement quality in different platforms being strong predictors of detection of variant *cis-*effects. We therefore leveraged predictions from the LoF and missense models to identify a set of high-quality *cis-*protein targets for which homozygous pLoF and missense variants were predicted to impact plasma abundance. Indeed, the detection rate of *cis-*protein under- or over-expression among this set of likely well measured targets was much higher (75% of homozygous pLoFs and 11% of homozygous missense variants) (**Supplementary Fig.11**).

## Discussion

Here, we leveraged broad-capture plasma proteomics in an underrepresented population with high rates of consanguinity and autozygosity to characterise the molecular and clinical consequences of rare homozygous LoF and missense variants (i.e. human knockouts). We showed the value of a ‘bottom-up’ approach in which systematic clinical characterisation of protein-informed human knockouts can enable 1) identification of individuals with likely undiagnosed rare disease characteristics, 2) discovery of novel disease-causing variants and 3) discovery of unreported gene-phenotype associations in humans. Of note, very few of the poorly characterised (e.g. amber or red in PanelApp) or novel gene-phenotype links we robustly identify here, have been described through recent extremely large-scale exome sequencing data ^36^. This highlights the power of bottom-up approaches for systematic characterisation of rare disorder gene-phenotype links.

For protein-informed rare homozygous LoF and missense variants, we saw extreme effect on *cis-*protein abundance ranging from differences in ∼20 standard deviations lower to 30 standard deviations higher than the average in the population. Integration of 3 separate proteomic platforms showed the majority (∼62%) of LoF and ∼32% of missense effects were consistent across 2 or more platforms. Integration of phenotypic information and predictions from machine learning models we developed using “gold-standard” findings (i.e. LoF or missense variants with evidence of an effect on cis-protein abundance from 2 or more platforms), suggested that at least ∼1/3 of platform specific variant effects were likely to represent true positives rather than epitope effects and other technical artifacts. Discrepancies between platforms can likely point to targeting of different isoforms, presence of epitope effects for specific platforms or a lack of assay specificity (that is, affinity reagent cross-reactivity with other proteins or lack of peptide-to-protein mapping specificity in MS). Systematically characterisation of variant, gene and protein characteristics associated with detection of *cis-*protein effects, showed variant quality control metrics (indicating likely sequencing artifacts rather than true variants) and pathogenicity scores as important predictors explaining the lack of effects on *cis-*protein abundance. However, the strongest predictors were proxies of protein assay quality such as the detectability in plasma or cross-platform correlations. This strongly points towards the need to develop and test more sensitive proteomic assays that can detect even low protein concentrations in blood to improve identification of rare variant effects. This was supported by detection rates of rare homozygous pLoF and missense variant *cis-*effects being substantially higher among well-measured proteins.

We further showed the potential for compensatory mechanisms ^40^ from other rare and common variants in the affected gene or influencing the protein proximally or distally (*cis-* or *trans-*pQTLs) to counteract strong effects from homozygous LoF or missense variants for selected examples. For example, we demonstrated the importance of common pQTL scores to explain the lack of an observed effect of homozygous pLoF of missense variants on *cis-* protein abundance, highlighting the interplay between common and rare variants in modulating a molecular or clinical phenotype as has been described for neurodevelopmental disorders previously ^41,42^.

Our study had several strengths, such as being the first to systematically characterise the proteomic and clinical impact in human knockouts through a bottom-up approach in an underrepresented population that is at high-risk given the high rate of autozygosity ^43^. The use of three separate proteomic platforms enabled categorisation of our findings into different tiers of strength of evidence. However, there are several limitations worth highlighting. While broad-capture proteomic technologies are revolutionising discovery in population-based studies, the can only inform on abundance but not on function. Second, while the use of EHRs provides a readily accessible and widely available strategy for phenotypic characterisation, this can lead to biased or incomplete phenotyping and uncertainty on the age of symptoms of disease onset. Furthermore, the common disease ICD-10 and SNOMED nomenclatures used in hospital episode statistics and primary care is poorly suited for rare disease phenotyping, which mostly relies on the human phenotype ontology. While previous work has pioneered mapping common and rare disease terminology ^44,45^, future work should focus on improving integration between common and rare disease nomenclature. Furthermore, the relatively small sample size meant we mostly only identified one carrier for each rare homozygous pLoF or missense variant. Future larger-scale efforts could improve molecular and phenotypic characterisation in multiple human knockouts with the exact same variant.

Overall, our study highlights the value of proteome guided characterisation of human knockouts to improve our understanding on the fundamental biology of gene functions and clinical characterisation and molecular diagnostics of undiagnosed rare disease patients.

## Methods

### Study design

Genes & Health (G&H) is a cohort of over 65,000 South Asian (Bangladeshi and Pakistani) individuals living in the United Kingdom (UK). Inclusion criteria included individuals aged 16 and over, from self-reported Bangladeshi and Pakistani backgrounds. Recruitment has been ongoing since 2015. The baseline assessment was performed at recruitment, upon which participants filled in a brief questionnaire, provided a saliva sample for genotyping and sequencing, and consented to being recalled and to longitudinal linkage to electronic health records (EHR) including data from primary, secondary care, cancer death registry. Details of the cohort have been previously described^15^. G&H was approved by the London Southeast NRES Committee of the Health Research Authority (14/LO/1240).

The current study comprises a subset of 1569 participants from the G&H cohort that were recalled for additional phenotyping and blood sampling. Individuals were selected for recall based on identification of genotypes of interest (i.e. participants without homozygous pLoF variants) and additional controls, enriching this subsample for human knockouts for a diverse set of cardiometabolic and immune genes. EDTA-plasma (non-fasted or random) samples were collected and processed according to SomaLogic’s recommended protocol ^46^, which involved storing processed samples at −80°C in under 1 hour.

### Whole Exome Sequencing

The Broad Institute performed ‘Standard Germline Exome v6’ using Twist exome capture reagents and Illumina 150bp PE Novaseq 6000 sequencing. BWA-MEM was used to map to the reference genome hg38 with ALT contigs to produce single individual gVCF and cram output files. Preprocessing and variant calling was performed using the Exome Germline Single Sample 3.0.0 pipeline^47^ using Picard 2.23.8^48^, GATK 4.2.2.0 HaplotypeCaller^49^, and Samtools 1.11 ^50^. The Broad Institute performed basic quality control statistics and delivered crams with >85% bases at >20X Twist bait target coverage. Chromosome Y and MT variants were not called, and chromosome X variants were called diploid for females and males.

Sample quality control (QC) was applied to remove those with <85% bases at >20X coverage in Gencode exons; with the contamination estimate freemix >0.03; self-stated gender that did not match biological sex inferred from exome data (and could not be reconciled); without a valid NHS number, sample duplicates (the lowest coverage sample(s) were removed). Joint genotype calling was performed on these samples using HAIL and GATK GenotypeGVCFs using the Broad Institute Joint Genotyping pipeline ^47^. WES crams were compared to 44,396 Illumina GSAv3 chip genotyping samples by using 3,596 common (MAF>0.001) SNPs that are in both WES and GSA hard called genotypes (without imputation) to identify high confidence matches. WES samples with mismatches were removed after comparing them to the GSA data as likely recruitment or laboratory errors. We further removed samples not predicted to be of South Asian ancestry based on PCA and reference samples from the 1000 Genomes Project. Individuals were stratified into Bangladeshi, Pakistani and other South Asian groups based on PCA. Samples were further excluded if the number of SNVs, transition/transversion ratio, number of transitions, number of transversions, number of insertions, number of deletions, insertion/deletion ratio were outside the median ±6 median absolute deviations (MAD) compared to samples from the same population, or if the heterozygote/homozygote ratio, heterozygosity rate was higher than the median + 6 MADs (to avoid removing samples with high autozygosity).

### Variant-level quality control

A random forest model was trained on chromosome 20 using the true positive (high confidence variant sites discovered in the 1,000 Genomes Project, SNVs present on the Illumina Omni 2.5 genotyping array and found in the 1,000 Genomes Project^51^, INDELs present in the Mills and Devine data^52^, HapMap3^53^ SNVs and INDELs) and false positive variants (QD<2 OR FS>60 OR MQ<30) as described above and then applied to the whole dataset. Features selected included quality by depth (QD), mean heterozygous allele balance, multiallelic site, strand odds ratio (SOR), mapping quality (MQ), number of alternative alleles at a site, variant type, site for which alleles include a ‘*’ allele, rank sum test for mapping qualities of reference versus alternative reads, multiallelic site containing SNVs and indels, rank sum test for relative positioning of reference versus alternative alleles within reads and allele type. Variants were ranked by their random forest score (i.e. the probability a variant reflects a true positive) and binned. We selected a random forest bin of 77 (true positive rate = 98.76%, false negative rate = 0.70%) and 57 (true positive rate = 95.65%, false negative rate = 4.88 %) to retain SNVs and indels, respectively.

### Genotype-level quality control

Among variants passing variant QC criteria, we further tested a combination of random forest bin, depth (DP), genotype quality (GQ), heterozygous allele balance (hetAB) and call rate to perform genotype-level QC. Specifically for variants passing the given random forest bin, genotypes were set to missing if they did not pass one or more of the listed genotype QC criteria. To determine the optimal combination, we calculated the percentage of true and false positives, the transmitted / untransmitted synonymous singleton ratio in trios (inferred using KING ^54^), the total number of Mendelian errors in trios and the mean number of heterozygous calls in runs of homozygosity (ROHs), for each combination of QC filters. For X chromosome variants, DP and GQ thresholds were tested separately for males and females and we calculated the mean number of heterozygous calls in non-pseudoautosomal regions in males instead of number of heterozygous calls in ROHs. The final set of filters selected were random forest bin 80, DP 10, GQ 20, hetAB 0.2 for SNPs, and random forest bin 80, DP 10, GQ 20, hetAB 0.2 and call rate 0.95 for SNVs; and random forest bin 44, DP 10, GQ 20, hetAB 0.3 and call rate 0.95 for indels.

### Proteomic Profiling

Proteomic profiling was performed from plasma samples using three different technologies: the Olink Explore HT platform (targeting 5416 unique proteins by 5420 assays), SomaScan 11k v5 (targeting 9664 unique proteins by 11083 aptamers) and the Seer Proteograph platform (which detected 8079 protein groups for 7762 unique proteins in at least 1 sample).

#### Olink Explore HT

Proteomic profiling with the Olink Explore HT platform was performed in 1641 samples from 1447 individuals. Details of the Olink platform have been previously described in detail^55^. Briefly, Olink relies on proximity extension assays, which targets proteins by pairs of antibodies conjugated to complimentary oligonucleotides. Protein assays are grouped across eight dilutions blocks, with blocks 1 to 4 having a 1:1 dilution (i.e. including low abundant proteins in plasma), block 5 a 1:10 dilution, block 6 a 1:100 dilution, block 7 a 1:1,000 dilution and block 8 a 1:100,000 dilution (i.e. including the most abundant proteins in plasma). Upon binding to their target protein, hybridization between probes enables amplification and subsequent relative quantification through next generation sequencing. Olink’s internal controls involve an incubation (a non-human antigen with matching antibodies), extension (IgG conjugated with a matching oligo pair) and amplification controls (synthetic double stranded DNA). Additional external controls are included in each plate, namely negative, plate (increased to 5 compared to the 3 included in their previous version of the Explore platform) and sample controls. We calculated limits of detection for each protein assay per plate based on negative controls run in duplicate. Normalised protein expression (NPX) values are generated by normalisation to the extension control, log2 transformation and further normalisation to the plate controls. Samples are flagged as a FAIL (and no NPX is calculated) if there are < 10k read counts per sample, or if incubation, extension or amplification controls have <150 counts per sample. Blocks are flagged as FAILs (and no NPX values are calculated) if <3 plate controls or <1 negative control pass quality control. We excluded 19 samples due to 1) >50% of protein assays failed or 2) >50% of protein assays with counts below the average count in negative control samples. We assessed assay reproducibility in one sample that was included in all plates (median spearman pairwise correlations between plates = 0.74 (range: 0.68 – 0.77)). We kept only the first sample taken for individuals with more than one sample and excluded individuals with no linkage to EHRs (N=1), leaving 1444 individuals for analysis.

#### SomaScan 11k v5

Proteomic profiling with the SomaScan 11k v5 platform was performed in 1751 samples from 1555 individuals. Details of the SomaScan platform have been previously described in detail ^56^. Briefly, SomaScan relies on modified DNA-based aptamers that recognise their target protein. Aptamers are grouped across 3 dilutions bins: 20% (1:5), 0.5% (1:200) and 0.005% (1:20,000). SomaLogic’s workflow includes the generation of scaling factors to account for intra- and inter-plate effects, for a hybridization normalisation, median signal normalisation, plate-scale normalisation and interplate calibration steps. Proteins are quantified with relative fluorescence units (RFUs) and finally normalised Adaptive Normalisation by Maximum Likelihood (ANML). SomaLogic does not provide LOD values. We excluded 4 samples that were strong outliers with a median sample RFU more than 3 standard deviation away from the average of the population. We assessed assay reproducibility in one sample that was included in all plates (median spearman pairwise correlations between plates = 0.99 (range: 0.98 – 0.99)). We kept only the first sample taken for individuals with more than one sample and excluded individuals with no linkage to EHRs (N=1), leaving 1444 individuals for analysis.

#### Seer Proteograph

We performed proteomic profiling in 1612 plasma samples from 1464 individuals using the Proteograph XT Assay^57,58^. Proteins were quantitatively captured via nanoparticle (NP)- associated protein coronas, then denatured, reduced, alkylated, and enzymatically digested with Trypsin and LysC. Resulting peptides were purified, quantified, vacuum-dried overnight, and reconstituted at 50 ng/µL.

Peptides (400 ng per injection; 8 µL) underwent Data-Independent Acquisition (DIA) analysis using a Vanquish NEO nanoLC coupled to an Orbitrap Astral mass spectrometer (Thermo Fisher). Peptides were separated using a trap-and-elute configuration (Acclaim PepMap 100 C18 trap column; 50 cm µPAC analytical column) at a flow rate of 1 µL/min over a 22-min gradient from 5–25% solvent B (0.1% FA in ACN), totalling a 33-min run. Mass spectrometry employed MS1 scans (380–980 m/z; Orbitrap detector; resolution 240,000; 0.6 s cycle; 500,000 ion AGC) and 200 fixed-window MS2 DIA scans (150–2000 m/z; 3 Th isolation windows; Astral detector; 25% collision energy; 50,000 ion AGC).

DIA mass spectrometry data were analyzed using the Proteograph™ Analysis Suite’s cloud pipelines^59^, employing the DIA-NN search engine (v1.8.1) ^60^ in library-free, match-between-runs mode. MS/MS spectra were matched against an in silico-generated spectral library based on human protein entries (UniProt UP000005640_9606), incorporating cohort-specific protein variants with minor allele count (MAC) ≥5. Search parameters included trypsin digestion (one missed cleavage), N-terminal methionine excision, fixed cysteine carbamidomethylation, peptide length 7–30 amino acids, precursor mass range 300–1800 m/z, fragment ion range 200–1800 m/z, and mass accuracy set to 3 ppm (MS1) and 8 ppm (MS2). Precursor and protein group FDR thresholds were set at 1%. Precursor-level data were annotated as "plus" (all variations retained), "minus" (all variable sites purged), or "reference" (only alternate variants purged), following Suhre et al. 2024^61^. Libraries were filtered at MAC ≥5 and minor allele frequency (MAF) ≥1%. We used the “reference” precursor data for all analyses.

Raw precursor intensities from DIA-NN were normalized using external synthetic peptide standards (PepCal) spiked into samples to account for LC-MS instrument drift. PepCal peptides were quantified via targeted extraction, quality-filtered based on retention time, intensity variation (CV), and completeness, and converted into fold-change relative to median peptide intensities across all runs. Normalization factors were calculated using the median fold-change across high-quality PepCal peptides, further smoothed over every five consecutive runs. Normalized precursor intensities per (biosample, nanoparticle) pair were aggregated into protein-level intensities for each biosample using the MaxLFQ algorithm (*fast_MaxLFQ*, R *iq* package^62^), minimizing variance in protein group intensities. Protein-level intensities were batch-corrected at the Proteograph plate level using *limma*’s *removeBatchEffect* function. Following batch correction, we identified a total of 8079 protein groups seen in at least one sample, with 5768 protein groups seen in ≥70% of the samples. Based on a blood sample measured repeatedly on each plate, we observed a median coefficient of variation of 22.8% (IQR: 17.8-30.6%) across 7089 protein groups seen at least 5 times. We subsequently performed principal component analysis on the batch corrected aggregated and log2-transformed protein groups that had less than 10% missing values. We identified a total of 67 samples that significantly differed (p-value<3.1×10^-5^) from a multi-dimensional normal distribution based on the first four principal components based on the Mahalanobis distance. Those samples were subsequently excluded from downstream analysis and manual inspection confirmed potential measurement issues rather than rare genetic effects as a source of deviation. For samples measured multiple times, we took only the first one measured leaving a total of 1429 samples for downstream analysis.

### Identification of human knockouts

Variants were annotated using the Ensembl Variant Effect Predictor (VEP version 105 with LOFTEE v1.04_GRCh38^63^) as specified by the BRAVA Consortium. We used only the MANE (Matched Annotation from NCBI and EBI) Select transcripts^64^, which are the primary transcripts used in the gnomAD browser. We identified all candidate human knockouts as those with 1) homozygous variants causing frameshift, stop-gain or affecting splice acceptor or donor sites in the MANE transcript (pLoF high or low confidence by LOFTEE) with a MAF < 1% and 2) homozygous missense variants with a MAF < 1%. We compared demographic characteristics between candidate human knockouts against individuals without any homozygous LoF or missense variants and against the entire G&H study population (**Supplementary Table 1**).

### Protein outlier detection

For each candidate “human gene knockout” we analysed each protein to compare its abundance in plasma relative to the distribution in all other individuals. We generated residual protein values by regressing out age, sex and the first 3 proteomic principal components to account for technical variation. We scaled the residual protein values to generate z-scores. To estimate p-values we computed the value of the cumulative density function of the normal distribution with the observed mean and standard deviation for each of the proteins. We adjusted p-values applying a Bonferroni correction to account to multiple comparisons among all protein assays within each patient (5420 for Olink, 11083 for SomaLogic, and 7762 for Seer). We first defined outliers in each platform as those with and absolute z-score > 3 and a bonferroni adjusted p-value <0.05. For Seer, we further performed a peptide-based analysis which compared the number of peptides mapping to the *cis-*protein detected in the homozygous pLoF or missense carrier against the rest of the population. We defined Seer outliers as those with an absolute z-score > 3 or those with a significantly lower peptide count for proteins that were detected in at least 70% of the population and more than 5 mapped peptides on average in the population. To determine cross-platform generalisability, we took protein outliers identified in each separate platform and assessed whether the absolute z-score was at least > 2 in the other platforms or had a significantly lower peptide count in Seer (for proteins detected in at least 70% of the population and more than 5 mapped peptides on average in the population).

### Phenotype matching scores and proxies of disease burden

We first identified HPO terms present in each human knockout by mapping ICD-10 4 digit codes to phecodes (89.86% of ICD-10 codes present in at least one human knockout were mapped to at least one phecode), and phecodes to HPO (60.31% of ICD-10 mapped phecodes were mapped to at least one HPO term) terms using maps previously developed and implemented in the R package phers (v1.0.2)^65^. Phenotype similarity scores were calculated as a semantic similarity score between HPO terms associated to the target gene (carrying the homozygous pLoF or missense variants) and HPO terms present in the human knockout for the target gene. We implemented this analysis using the R package PCAN (v.1.30.0)^20^. We normalised all generated phenotype similarity scores to the highest score obtained in any of the individuals carrying a homozygous LoF or missense variant included in the proteomic study. We considered human knockouts to match phenotypically to expected phenotypes if phenotype similarity score ≥ 0.2. We determined this threshold by manual chart review of human knockouts for green genes in PanelApp.

We further computed the number of unique diagnoses in each individual as the number of ICD-10 4-digit codes. This included diagnoses from primary care that were mapped from SNOMED-CT to ICD-10. We then identified human knockouts with a significantly higher (p-value <0.05) number of diagnoses compared to the individuals without homozygous missense or pLoF variants. For this we generated p-values by computing the cumulative density function of the normal distribution with the observed mean and standard deviation for the number of ICD-10 4-digit codes. For these individuals we group diagnoses by ICD-10 chapter to identify organ systems most affected.

### Single-cell RNA sequencing expression

We downloaded single-cell RNA sequencing data from the Human Protein Atlas ^35^ to generate expression (normalised transcripts per million) profile plots for genes of interest. Data was downloaded from https://www.proteinatlas.org/humanproteome/single+cell/single+cell+type/data#cell_type_data on the 18^th^ March 2025.

### Variant, gene and protein characteristics associated with detection of homozygous LoF and missense variant effect on *cis-*protein abundance

We curated a comprehensive list of 246 variables across 8 gene, variant and protein biological and technical characteristic groups (**Supplementary Table 6**). Variant level features were obtained from VEP and VEP plugins, individual level WES quality/call data and Repeat Masker (https://repeatmasker.org/), pext scores were obtained from gnomAD v.1 release^63^. Gene and protein characteristic were retrieved from UniProt^66^. Protein assay quality characteristics were obtained from data directly provided by Seer, SomaScan or Olink or derived from it. We ran logistic regression models to test the association of each variable with detection of *cis-* protein under- or over-expression as a binary variable. Categorical variables were one-hot encoded. Continuous variables with strong deviations from a normal distribution were log transformed and all continuous variables were scaled to a mean of zero and standard deviation of 1. We restricted this analysis to variants for which observed no effect on cis-protein abundance and those for which we observed cis-protein under- or over-expression with at least 2 different proteomic platforms. Variants for which we only observed an effect with 1 proteomic platform were excluded to minimize the risk of including epitope effects. We further ran logistic regression model for each of the variable tested while adjusting for additional covariates known to affect the efficiency of nonsense mediated RNA decay (MAF, position in the coding sequence, protein position, and number of exon in which the variant was located) and for whether the variant was a pLoF or missense. We ran three type of analysis including 1) all pLoF and missense variants, 2) missense only of 3) pLoF only variants. For the missense only analysis, we restricted to 224 variables, as we excluded variables that were specific to pLoF variables only or not applicable to missense variants (e.g. intron position, LOFTEE high-confidence or low confidence annotation, etc.). Similarly for pLoF only analyses we restricted to 218 variables after excluding those only applicable to missense variables or that had only one class recoded among pLoFs.

### Metabolome profiling and metabolite outlier detection

Metabolite measurements were performed using the Metabolon HD4 untargeted platform in EDTA plasma samples for 1538 individuals from the GCH cohort who had also undergone proteomic profiling with at least one platform. The Metabolon platform covers a range of metabolites across biochemical groups, including amino acids, energy molecules, carbohydrates, lipids, peptides, nucleotides and carbohydrates, as well as uncharacterized, but reliably captured metabolites. We analysed 1148 metabolites that were detected in at least 70% of the study population.

Metabolite outliers were generated similar to the proteome, by converting measurements to Z-scores. We adjusted the resulting p-values across the number of metabolites tested and considered metabolites significant outliers when p < 0.05/1148.

### Predicting detection of homozygous LoF and missense variant effect on *cis-*protein abundance

We developed a machine learning model using extreme gradient boosting (XGBoost) to predict the likelihood of a rare homozygous pLoF or missense variants having an effect on *cis-* protein plasma abundance, and to identify important features to make these predictions. We chose XGBoost given its ability to handle mixed class variables more efficiently than other models. In the same way as for the association analyses, we developed three models for: 1) pLoF and missense variants (LoF+missense) using 240 variables as predictors, 2) missense variants only using 224 variables as predictors and 3) pLoF variants only using 218 variables as predictors. We further trained two additional pLoF+missense models by using genetic only predictors (i.e. excluding variables categorised under the “protein assay quality” and “protein characteristics” groups) or protein only predictors (i.e. excluding variables categorised under the “variant pathogenicity”, “variant QC”, “polygenic background”, “variant / gene characteristics” and “gene-phenotype association” groups). We restricted this analysis to variants for which observed no effect on cis-protein abundance and those for which we observed cis-protein under- or over-expression with at least 2 different proteomic platforms, which we split into 75% for training and 25% for validation. In the training set, we performed a grid search to optimise hyperparameters through 5-fold cross-validation. We optimised the following parameters through the following values: the learning rate or eta (0.01, 0.1, 0.3), maximum depth of a tree (4, 6, 8), subsample ratio of the training instances (0.7, 0.8, 0.9), subsample ratio of columns when constructing each tree (0.7, 0.8, 0.9). We selected the optimal parameter combination as the one leading to the minimum root mean square error during cross-validation, to train the final model. We extracted variable importance metrics from the final model. We used the R package xgboost v.1.7.8.1 to derive these models. We then applied the final model to the held-out validation set where we computed performance metrics including the AUROC, sensitivity and specificity. We selected a threshold that optimised a balance between both sensitivity and specificity using the R package pROC v1.18.0.

We further applied the final LoF+missense, LoF and missense models to the set of variants with evidence of *cis-*protein under- or over-expression detected through only one platform to predict the likelihood that these represent true positives according to our XGBoost models. We used the previously established thresholds which balanced sensitivity and specificity to categorise these platform specific findings into “predicted true positives” and “predicted not to impact *cis-*protein abundance”, the latter representing variants with the greatest potential of being epitope effects. Given the lack of a truth standard among the platform specific findings, we adopted two strategies to make inferences on sensitivity and specificity. 1) We used ClinVar benign variant annotations among this set to infer specificity of the predictions from our models. We were unable to use ClinVar pathogenic annotations to infer sensitivity as there were only 2 variants annotated as pathogenic in ClinVar in this set. 2) We used phenotype matching scores for these individuals, as those in which we observe a matching or expected phenotype (phenotype score ≥ 0.2) are more likely to represent true positives. We compared our models sensitivity and specificity among the set of platform specific predictions against established variant pathogenicity scores including: CADD^67^ (for both LoF and missense variants), BayesDel^68^ (for both LoF and missense variants), AlphaMissense^69^, REVEL, PolyPhen and SIFT^70^ (for missense variants only). For these scores we used established thresholds reported in the literature that are considered to identify pathogenic variants: ≥ 25 for CADD^71^, > 0.0692665 for BayesDel^71^, > 0.564 for AlphaMissense^69^, 0.5 for REVEL^72^, 0.908 for PolyPhen^73^ and < 0.05 for SIFT^70^.

## Acknowledgements

Genes & Health is/has recently been core-funded by Wellcome (WT102627, WT210561), the Medical Research Council (UK) (M009017, MR/X009777/1, MR/X009920/1), Higher Education Funding Council for England Catalyst, Barts Charity (845/1796), Health Data Research UK (for London substantive site), and research delivery support from the NHS National Institute for Health Research Clinical Research Network (North Thames). We acknowledge the support of the National Institute for Health and Care Research Barts Biomedical Research Centre (NIHR203330); a delivery partnership of Barts Health NHS Trust, Queen Mary University of London, St George’s University Hospitals NHS Foundation Trust and St George’s University of London

Genes & Health is/has recently been funded by Alnylam Pharmaceuticals, Genomics PLC; and a Life Sciences Industry Consortium of AstraZeneca PLC, Bristol-Myers Squibb Company, GlaxoSmithKline Research and Development Limited, Maze Therapeutics Inc, Merck Sharp & Dohme LLC, Novo Nordisk A/S, Pfizer Inc, Takeda Development Centre Americas Inc.

We thank Social Action for Health, Centre of The Cell, members of our Community Advisory Group, and staff who have recruited and collected data from volunteers. We thank the NIHR National Biosample Centre (UK Biocentre), the Social Genetic & Developmental Psychiatry Centre (King’s College London), Wellcome Sanger Institute, and Broad Institute for sample processing, genotyping, sequencing and variant annotation. This work uses data provided by patients and collected by the NHS as part of their care and support. This research utilised Queen Mary University of London’s Apocrita HPC facility, supported by QMUL Research-IT, http://doi.org/10.5281/zenodo.438045

We thank: Barts Health NHS Trust, NHS Clinical Commissioning Groups (City and Hackney, Waltham Forest, Tower Hamlets, Newham, Redbridge, Havering, Barking and Dagenham), East London NHS Foundation Trust, Bradford Teaching Hospitals NHS Foundation Trust, Public Health England (especially David Wyllie), Discovery Data Service/Endeavour Health Charitable Trust (especially David Stables), Voror Health Technologies Ltd (especially Sophie Don), NHS England (for what was NHS Digital) - for GDPR-compliant data sharing backed by individual written informed consent.

Most of all we thank all of the volunteers participating in Genes & Health.

A favourable ethical opinion for the main Genes & Health research study was granted by NRES Committee London - South East (reference 14/LO/1240) on 16 Sept 2014. Queen Mary University of London is the Sponsor, and Data Controller.

## Data availability

Individual-level data from Genes & Health are available for bona fide researchers on application (https://www.genesandhealth.org/).

## Code availability

Associated code will be deposited on GitHub upon final publication.

## References

1. Feuermann, M., et al. A compendium of human gene functions derived from evolutionary modelling. Nature 640, 146–154 (2025).

2. MacArthur, D.G., et al. A systematic survey of loss-of-function variants in human protein-coding genes. Science 335, 823–828 (2012).

3. Saleheen, D., et al. Human knockouts and phenotypic analysis in a cohort with a high rate of consanguinity. Nature 544, 235–239 (2017).

4. Spedicati, B., et al. Natural human knockouts and Mendelian disorders: deep phenotyping in Italian isolates. Eur J Hum Genet 29, 1272–1281 (2021).

5. Sulem, P., et al. Identification of a large set of rare complete human knockouts. Nat Genet 47, 448–452 (2015).

6. Narasimhan, V.M., et al. Health and population effects of rare gene knockouts in adult humans with related parents. Science 352, 474–477 (2016).

7. Fawzy, M. & Marsh, J.A. Understanding the heterogeneous performance of variant effect predictors across human protein-coding genes. Sci Rep 14, 26114 (2024).

8. Fowler, D.M. & Rehm, H.L. Will variants of uncertain significance still exist in 2030? Am J Hum Genet 111, 5–10 (2024).

9. Cummings, B.B., et al. Improving genetic diagnosis in Mendelian disease with transcriptome sequencing. Sci Transl Med 9(2017).

10. Gonorazky, H.D., et al. Expanding the Boundaries of RNA Sequencing as a Diagnostic Tool for Rare Mendelian Disease. The American Journal of Human Genetics 104, 466–483 (2019).

11. Yepez, V.A., et al. Clinical implementation of RNA sequencing for Mendelian disease diagnostics. Genome Med 14, 38 (2022).

12. Uhlen, M., et al. The human secretome. Sci Signal 12(2019).

13. Koprulu, M., et al. Proteogenomic links to human metabolic diseases. Nat Metab 5, 516–528 (2023).

14. Pietzner, M., et al. Mapping the proteo-genomic convergence of human diseases. Science 374, eabj1541 (2021).

15. Finer, S., et al. Cohort Profile: East London Genes & Health (ELGH), a community-based population genomics and health study in British Bangladeshi and British Pakistani people. Int J Epidemiol 49, 20–21i (2020).

16. Gerasimavicius, L., Liu, X. & Marsh, J.A. Identification of pathogenic missense mutations using protein stability predictors. Sci Rep 10, 15387 (2020).

17. Stefl, S., Nishi, H., Petukh, M., Panchenko, A.R. & Alexov, E. Molecular mechanisms of disease-causing missense mutations. J Mol Biol 425, 3919–3936 (2013).

18. Janes, J., et al. Predicted mechanistic impacts of human protein missense variants. bioRxiv (2024).

19. Lacoste, J., et al. Pervasive mislocalization of pathogenic coding variants underlying human disorders. Cell 187, 6725–6741 e6713 (2024).

20. Godard, P. & Page, M. PCAN: phenotype consensus analysis to support disease-gene association. BMC Bioinformatics 17, 518 (2016).

21. Online Mendelian Inheritance in Man, OMIM®. Vol. 2025 (McKusick-Nathans Institute of Genetic Medicine, Johns Hopkins University, Baltimore, MD).

22. Landrum, M.J., et al. ClinVar: public archive of relationships among sequence variation and human phenotype. Nucleic Acids Res 42, D980–985 (2014).

23. Martin, A.R., et al. PanelApp crowdsources expert knowledge to establish consensus diagnostic gene panels. Nat Genet 51, 1560–1565 (2019).

24. Yang, Y., et al. Clinical whole-exome sequencing for the diagnosis of mendelian disorders. N Engl J Med 369, 1502–1511 (2013).

25. Crawford, P.J., Aldred, M. & Bloch-Zupan, A. Amelogenesis imperfecta. Orphanet J Rare Dis 2, 17 (2007).

26. Chin, A., Balasubramanyam, S. & Davis, C.M. Very Elevated IgE, Atopy, and Severe Infection: A Genomics-Based Diagnostic Approach to a Spectrum of Diseases. Case Reports Immunol 2021, 2767012 (2021).

27. Investigators, S., et al. Darapladib for preventing ischemic events in stable coronary heart disease. N Engl J Med 370, 1702–1711 (2014).

28. Przybyszewska, J., Zlotogorski, A. & Ramot, Y. Re-evaluation of epidermodysplasia verruciformis: Reconciling more than 90 years of debate. J Am Acad Dermatol 76, 1161–1175 (2017).

29. Hegele, R.A., et al. Hepatic lipase deficiency. Clinical, biochemical, and molecular genetic characteristics. Arterioscler Thromb 13, 720–728 (1993).

30. Breckenridge, W.C., et al. Lipoprotein abnormalities associated with a familial deficiency of hepatic lipase. Atherosclerosis 45, 161–179 (1982).

31. Alnouri, F., et al. Xanthomas Can Be Misdiagnosed and Mistreated in Homozygous Familial Hypercholesterolemia Patients: A Call for Increased Awareness Among Dermatologists and Health Care Practitioners. Glob Heart 15, 19 (2020).

32. Zoodsma, M., et al. A genetic map of human metabolism across the allele frequency spectrum. medRxiv, 2025.2001.2030.25321073 (2025).

33. Groza, T., et al. The International Mouse Phenotyping Consortium: comprehensive knockout phenotyping underpinning the study of human disease. Nucleic Acids Res 51, D1038–D1045 (2023).

34. Aono, Y., et al. CD109 on Dendritic Cells Regulates Airway Hyperreactivity and Eosinophilic Airway Inflammation. Am J Respir Cell Mol Biol 68, 201–212 (2023).

35. Karlsson, M., et al. A single-cell type transcriptomics map of human tissues. Sci Adv 7(2021).

36. Jurgens, S.J., et al. Rare coding variant analysis for human diseases across biobanks and ancestries. Nat Genet 56, 1811–1820 (2024).

37. Hey, H.W.D., et al. Paraspinal myopathy-induced intervertebral disc degeneration and thoracolumbar kyphosis in TSC1mKO mice model-a preliminary study. Spine J 22, 483–494 (2022).

38. Chen, S., Young, M.F., Chakravarti, S. & Birk, D.E. Interclass small leucine-rich repeat proteoglycan interactions regulate collagen fibrillogenesis and corneal stromal assembly. Matrix Biol 35, 103–111 (2014).

39. Lindeboom, R.G., Supek, F. & Lehner, B. The rules and impact of nonsense-mediated mRNA decay in human cancers. Nat Genet 48, 1112–1118 (2016).

40. Gudmundsson, S., et al. Exploring penetrance of clinically relevant variants in over 800,000 humans from the Genome Aggregation Database. bioRxiv, 2024.2006.2012.593113 (2024).

41. Huang, Q.Q., et al. Dissecting the contribution of common variants to risk of rare neurodevelopmental conditions. medRxiv, 2024.2003.2005.24303772 (2024).

42. Pizzo, L., et al. Rare variants in the genetic background modulate cognitive and developmental phenotypes in individuals carrying disease-associated variants. Genet Med 21, 816–825 (2019).

43. Malawsky, D.S., et al. Influence of autozygosity on common disease risk across the phenotypic spectrum. Cell 186, 4514–4527 e4514 (2023).

44. Bastarache, L., et al. Improving the phenotype risk score as a scalable approach to identifying patients with Mendelian disease. J Am Med Inform Assoc 26, 1437–1447 (2019).

45. Bastarache, L., et al. Phenotype risk scores identify patients with unrecognized Mendelian disease patterns. Science 359, 1233–1239 (2018).

46. SomaLogic, I. The SomaScan Assay: Recommended Sample Handling and Processing for Core Sample Types. Vol. Rev 3 (2021).

47. Degatano, K., et al. WDL Analysis Research Pipelines: Cloud-Optimized Workflows for Biological Data Processing and Reproducible Analysis. in Preprints (Preprints, 2024).

48. Picard toolkit. (Broad Institute, Broad Institute, GitHub repository, 2019).

49. Poplin, R., et al. Scaling accurate genetic variant discovery to tens of thousands of samples. bioRxiv, 201178 (2018).

50. Danecek, P., et al. Twelve years of SAMtools and BCFtools. Gigascience 10(2021).

51. Genomes Project, C., et al. A global reference for human genetic variation. Nature 526, 68–74 (2015).

52. Mills, R.E., et al. An initial map of insertion and deletion (INDEL) variation in the human genome. Genome Res 16, 1182–1190 (2006).

53. International HapMap, C., et al. Integrating common and rare genetic variation in diverse human populations. Nature 467, 52–58 (2010).

54. Manichaikul, A., et al. Robust relationship inference in genome-wide association studies. Bioinformatics 26, 2867–2873 (2010).

55. Wik, L., et al. Proximity Extension Assay in Combination with Next-Generation Sequencing for High-throughput Proteome-wide Analysis. Mol Cell Proteomics 20, 100168 (2021).

56. Gold, L., et al. Aptamer-based multiplexed proteomic technology for biomarker discovery. PLoS One 5, e15004 (2010).

57. Blume, J.E., et al. Rapid, deep and precise profiling of the plasma proteome with multi-nanoparticle protein corona. Nat Commun 11, 3662 (2020).

58. Ferdosi, S., et al. Engineered nanoparticles enable deep proteomics studies at scale by leveraging tunable nano-bio interactions. Proc Natl Acad Sci U S A 119, e2106053119 (2022).

59. Guturu, H., et al. Cloud-Enabled Scalable Analysis of Large Proteomics Cohorts. J Proteome Res 24, 1462-1469 (2025).

60. Demichev, V., Messner, C.B., Vernardis, S.I., Lilley, K.S. & Ralser, M. DIA-NN: neural networks and interference correction enable deep proteome coverage in high throughput. Nat Methods 17, 41–44 (2020).

61. Suhre, K., et al. Nanoparticle enrichment mass-spectrometry proteomics identifies protein-altering variants for precise pQTL mapping. Nat Commun 15, 989 (2024).

62. Pham, T.V., Henneman, A.A. & Jimenez, C.R. iq: an R package to estimate relative protein abundances from ion quantification in DIA-MS-based proteomics. Bioinformatics 36, 2611–2613 (2020).

63. Karczewski, K.J., et al. The mutational constraint spectrum quantified from variation in 141,456 humans. Nature 581, 434–443 (2020).

64. Morales, J., et al. A joint NCBI and EMBL-EBI transcript set for clinical genomics and research. Nature 604, 310–315 (2022).

65. Aref, L., Bastarache, L. & Hughey, J.J. The phers R package: using phenotype risk scores based on electronic health records to study Mendelian disease and rare genetic variants. Bioinformatics 38, 4972–4974 (2022).

66. UniProt, C. UniProt: the Universal Protein Knowledgebase in 2025. Nucleic Acids Res 53, D609–D617 (2025).

67. Kircher, M., et al. A general framework for estimating the relative pathogenicity of human genetic variants. Nat Genet 46, 310–315 (2014).

68. Feng, B.J. PERCH: A Unified Framework for Disease Gene Prioritization. Hum Mutat 38, 243–251 (2017).

69. Cheng, J., et al. Accurate proteome-wide missense variant effect prediction with AlphaMissense. Science 381, eadg7492 (2023).

70. Ng, P.C. & Henikoff, S. SIFT: Predicting amino acid changes that affect protein function. Nucleic Acids Res 31, 3812–3814 (2003).

71. Pejaver, V., et al. Calibration of computational tools for missense variant pathogenicity classification and ClinGen recommendations for PP3/BP4 criteria. Am J Hum Genet 109, 2163–2177 (2022).

72. Ioannidis, N.M., et al. REVEL: An Ensemble Method for Predicting the Pathogenicity of Rare Missense Variants. Am J Hum Genet 99, 877–885 (2016).

73. Adzhubei, I.A., et al. A method and server for predicting damaging missense mutations. Nat Methods 7, 248–249 (2010).

