## Supplementary Figures for "Proteomic and clinical impact of human knockouts in British South Asians"

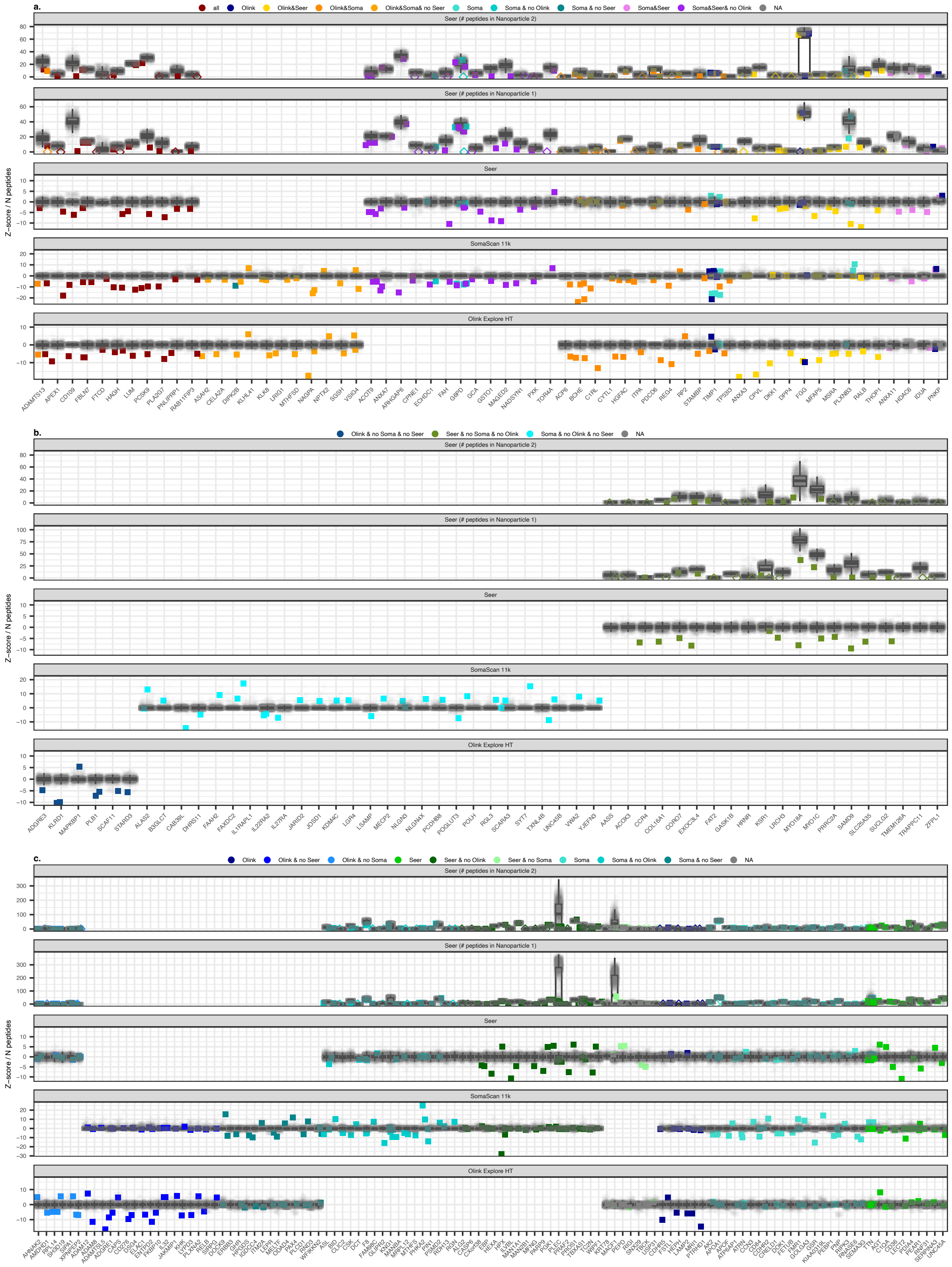


**Supplementary figure 1. Extreme effects of rare homozygous missense variants on *cis*-protein plasma abundance. a,** Missense variants with evidence of an effect on *cis*-protein abundance detected by at least one proteomic platform. Z-scores represent the number of standard deviations protein levels in the variant carrier are away from the population. For the Seer proteograph platform variant effects were further evaluated on the number of peptides detected in the variant carrier in comparison to the population distribution. Variants are categorised into **a)** those with evidence of a cis-protein effect detected by at least 2 platforms (>= platforms), **b)** those were the cis-protein is only covered by a single platform, and **c)** those covered by at least 2 different proteomic platforms but with evidence of an effect on the cis-protein detected by only one platform.

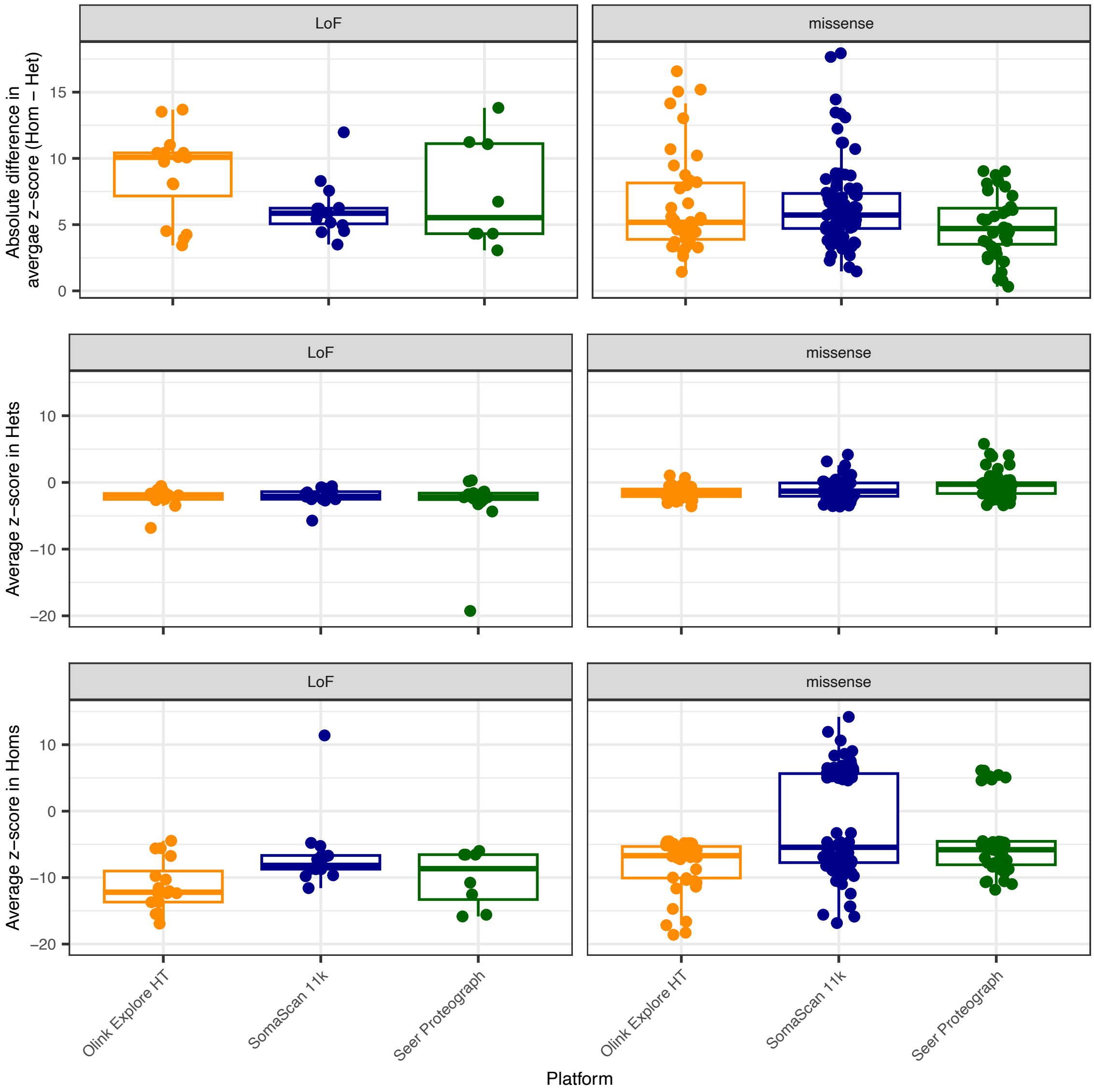


**Supplementary figure 2. Comparison of variant effects on *cis*-protein abundance in homozygous vs heterozygous pLoF or missense variant carriers.** Top: Absolute average difference in *cis*-protein z-scores between homozygous carriers and heterozygous carriers for pLoF and missense variants with evidence of an effect on the *cis*-protein abundance identified in each platform. Middle: Average *cis*-protein z-scores in heterozygous carriers for pLoF and missense variants. Bottom: Average *cis*-protein z-scores in homozygous carriers for pLoF and missense variants.

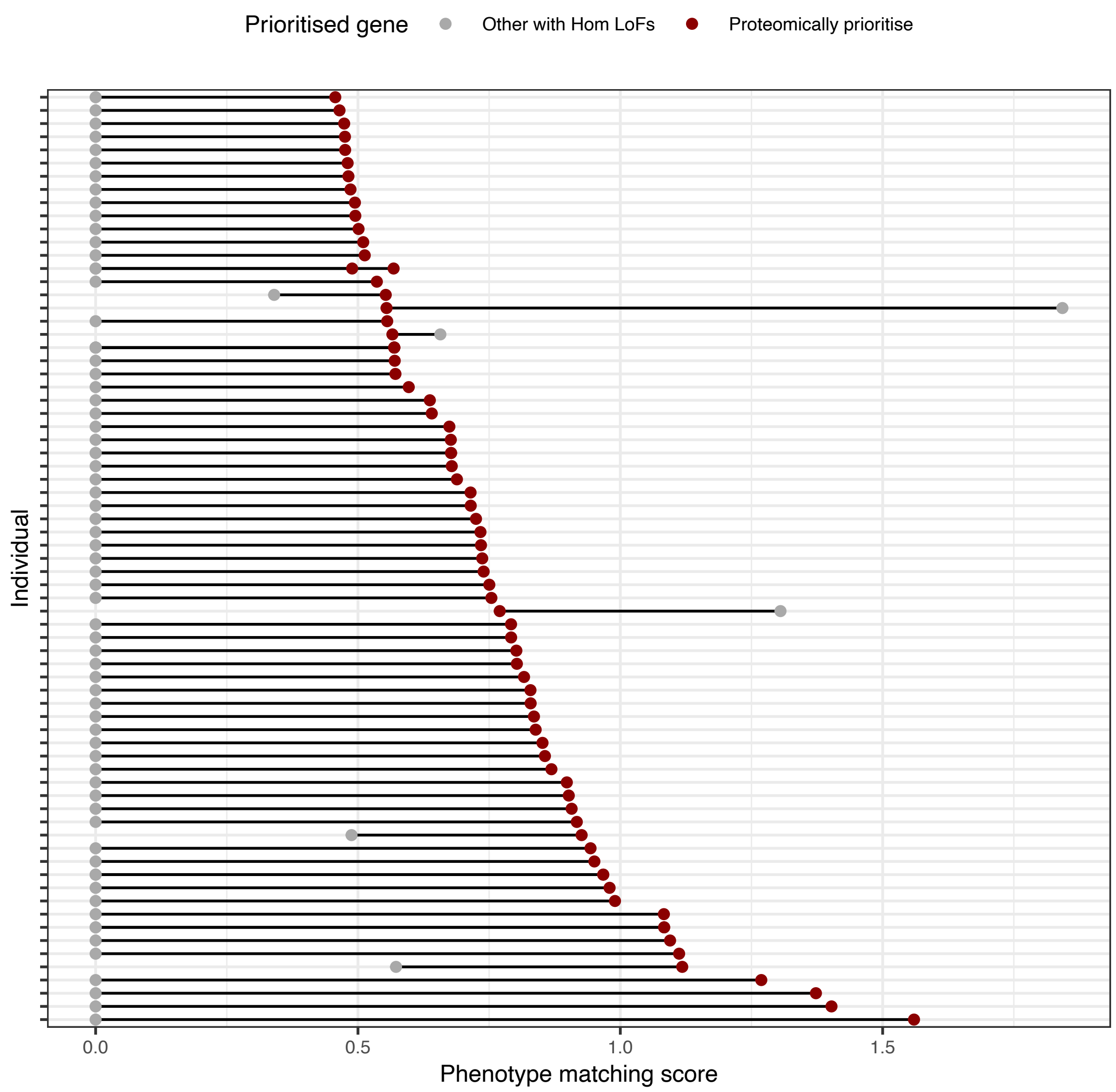


**Supplementary figure 3. Phenotype matching scores for protein-informed 'knockout' genes and any other gene with homozygous pLoF variants in the same individuals.** We compare the phenotype matching score for the gene with supporting evidence from proteomics (red) against phenotype matching scores for other genes with homozygous pLoF variants in the same individuals regardless of proteomic coverage (grey).

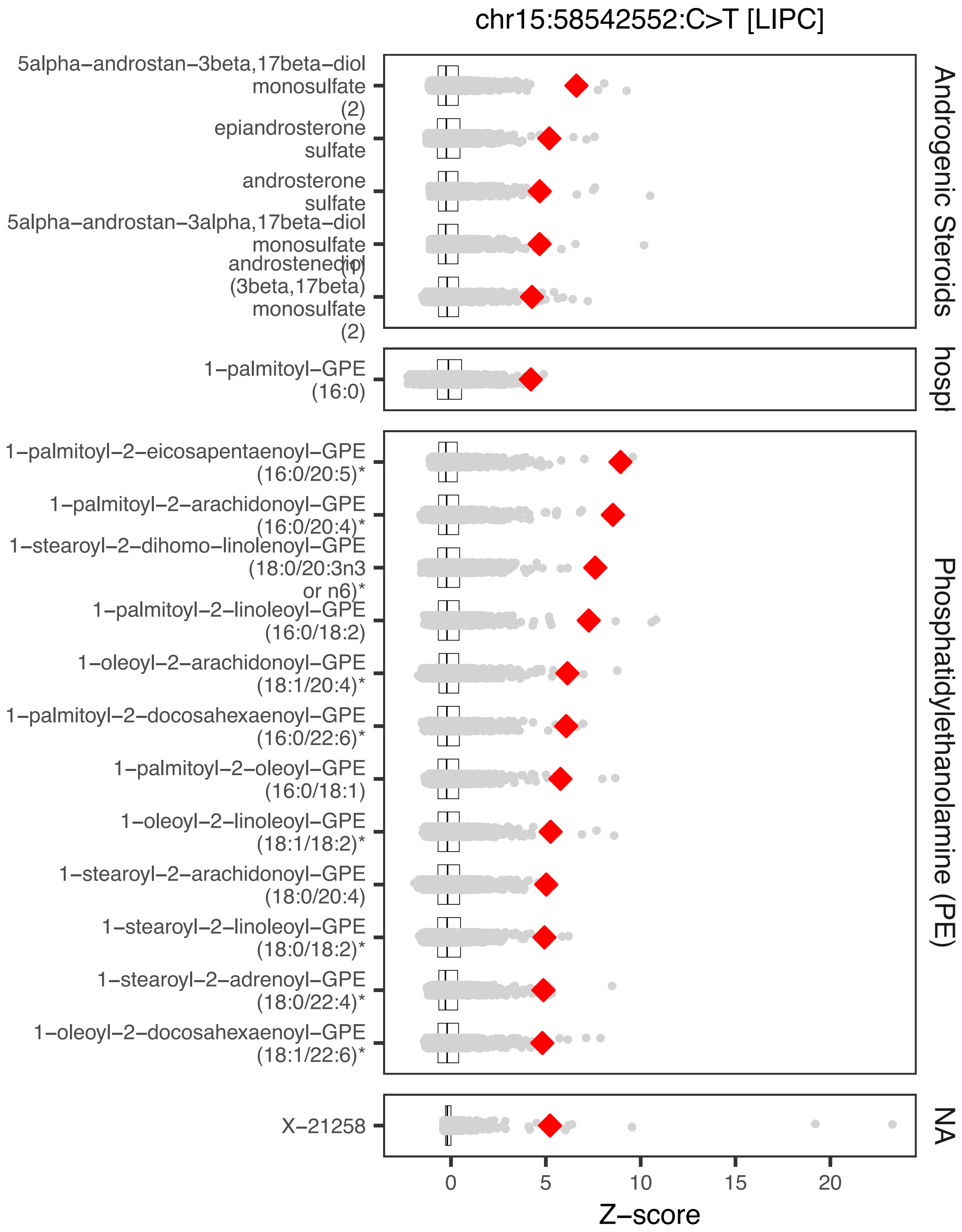


**Supplementary Figure 4. Metabolite outlier profile in an individual with a protein-informed knockout variant in the *LIPC* gene.** Metabolite z-scores for the LIPC knockout are shown in comparison to the remainder of the population. Only significant metabolite outliers identified out of the entire Metabolon HD4 platform are shown.

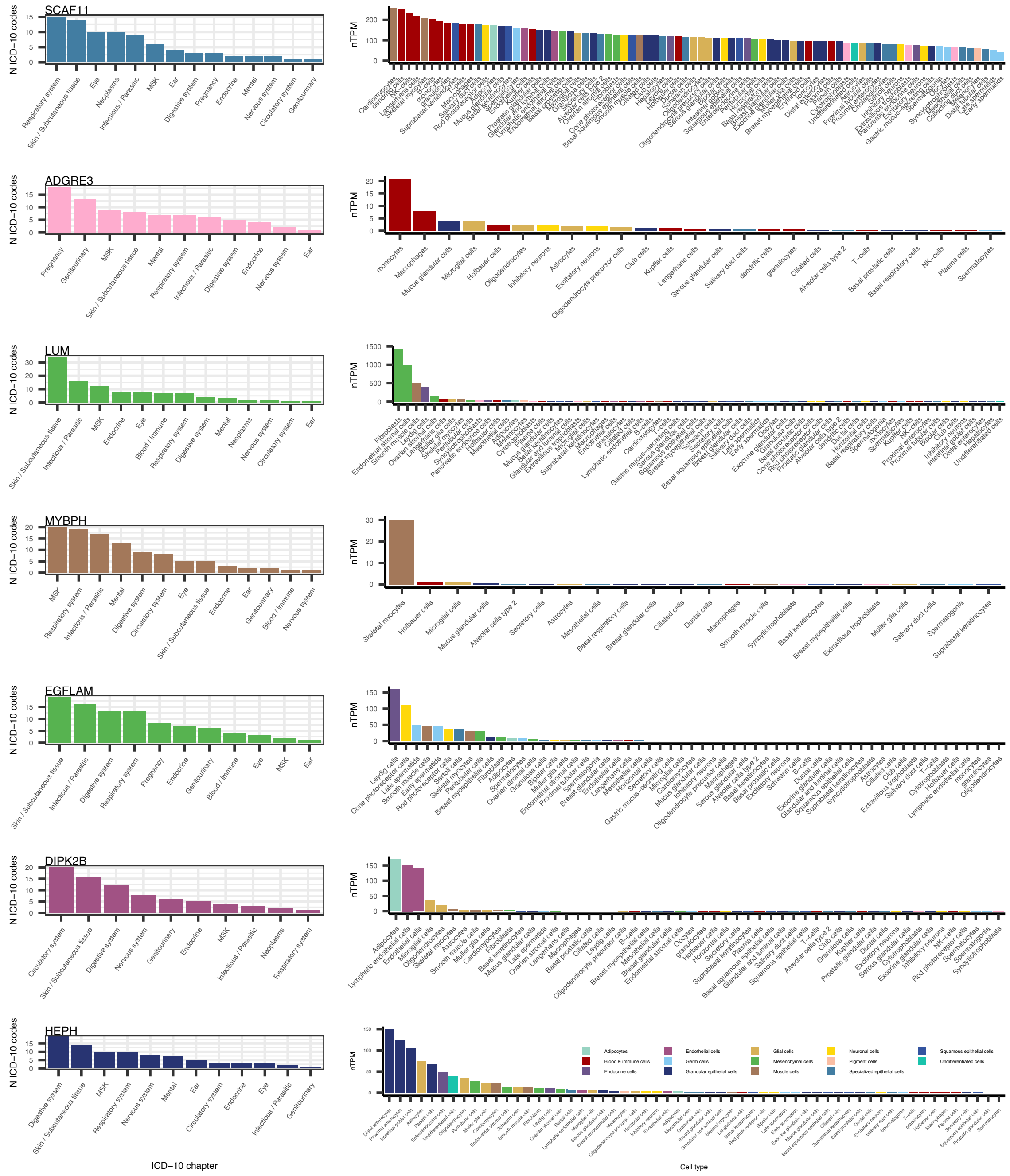


**Supplementary figure 5. Characterisation of clinical impact of human knockouts without a phenotype matching score. Left:** Number of unique diagnoses per ICD-10 chapter in protein-informed human knockouts in the 5% of the distribution of the number of diagnostic codes. **Right:** Ranked expression across cell types for target genes **(left)** from single cell RNAseq data from the Human protein Atlas.

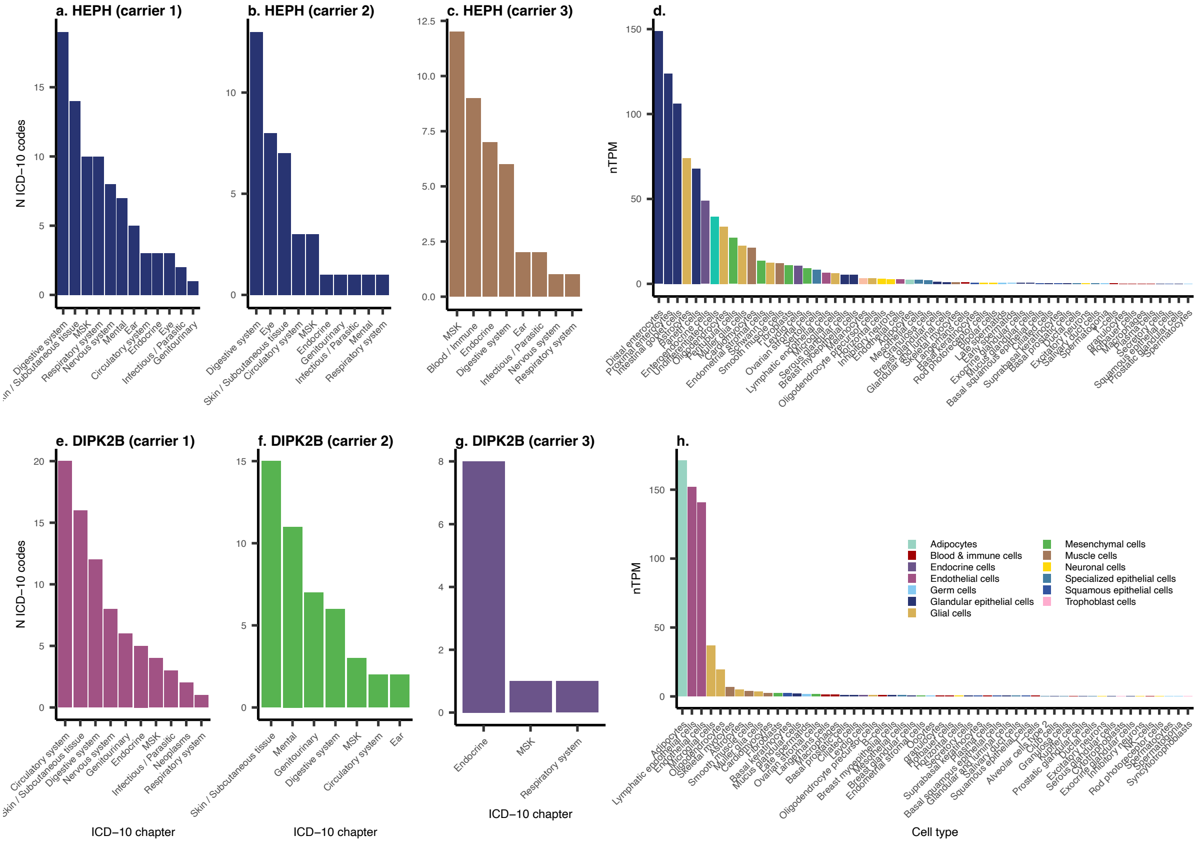


**Supplementary figure 6. Characterisation of clinical impact of human knockouts without a phenotype matching score.** Number of unique diagnoses per ICD-10 chapter in protein-informed human knockouts for **(a-c)** HEPH and **(e-g)** DIPK2B**.** Ranked expression across cell types for **(d)** HEPH and **(h)** DIPK2B from single cell RNAseq data from the Human protein Atlas.

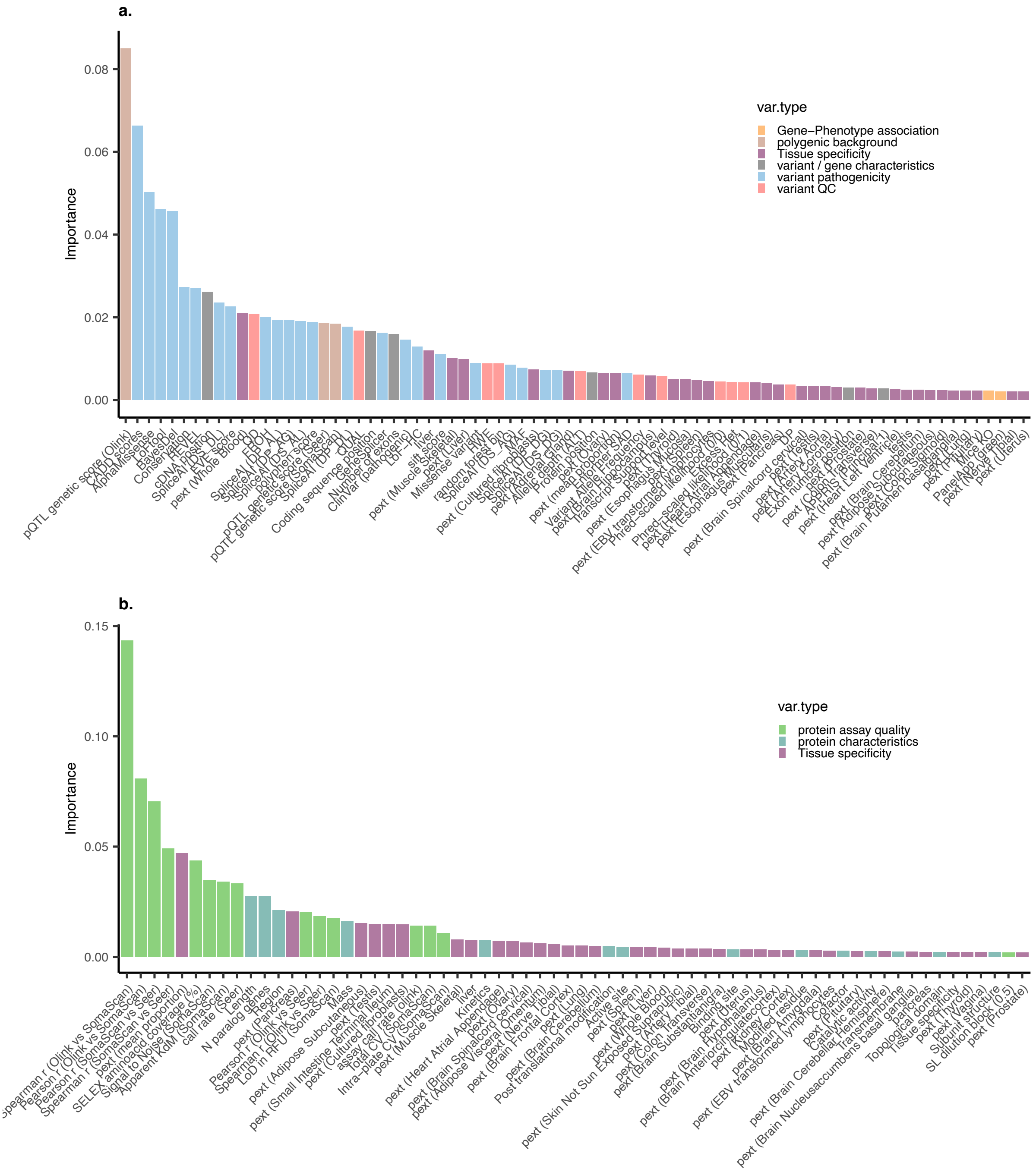


**Supplementary figure 7. Variable importance for XGBoost models trained to predict the likelihood of seeing and effect of rare homozygous LoF and missense variants on *cis*-protein abundance. a,** Variable important for models trained on genetic data only, that is, excluding all protein characteristics and assay quality. **b,** Variable importance for models trained on protein data only, that is, excluding all genetic characteristics.

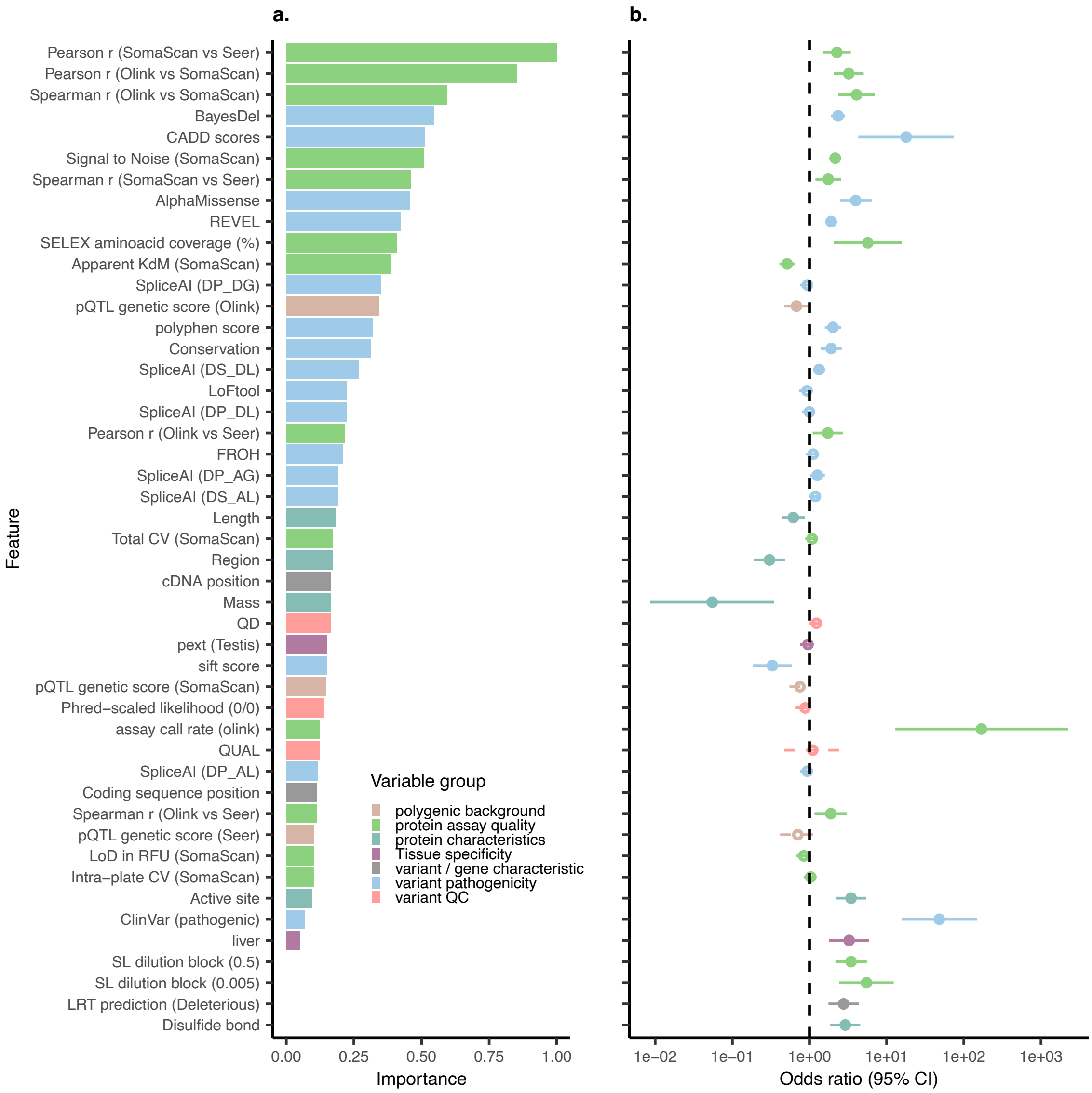


**Supplementary figure 8. Prediction of protein-informed homozygous missense human knockouts. a,** Ranked feature importance (normalised to the feature with the highest score) from an XGBoost model trained to predict detection of cis-protein outliers for homozygous missense variants (missense model). **b,** Associations between features and the likelihood of detecting a cis-protein outlier for all predictive feature or those significantly associated at a 5% FDR threshold.

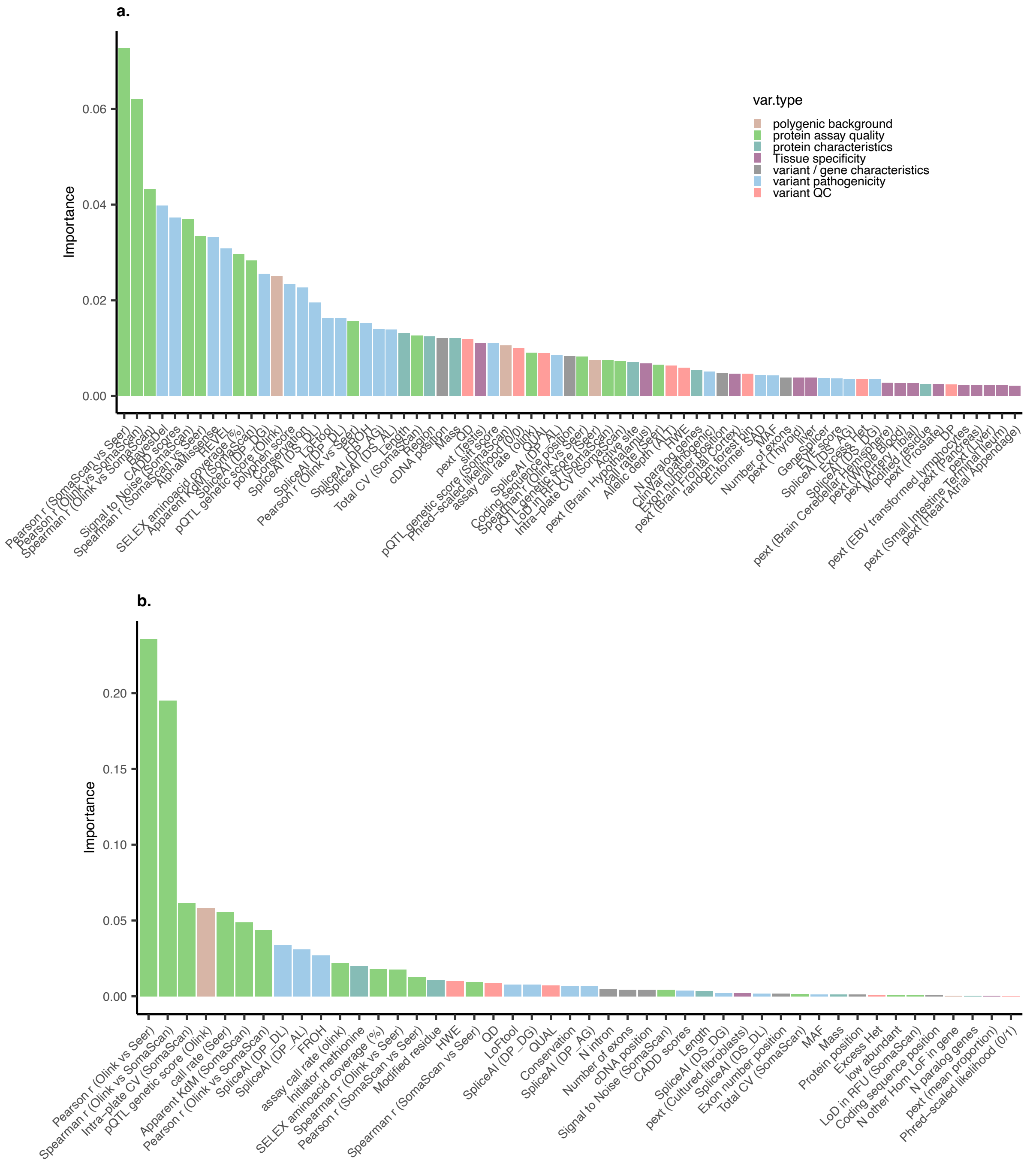


**Supplementary figure 9. Variable importance for XGBoost models trained to predict the likelihood of seeing and effect of (a) rare homozygous missense or (b) LoF variants on *cis*-protein abundance.**

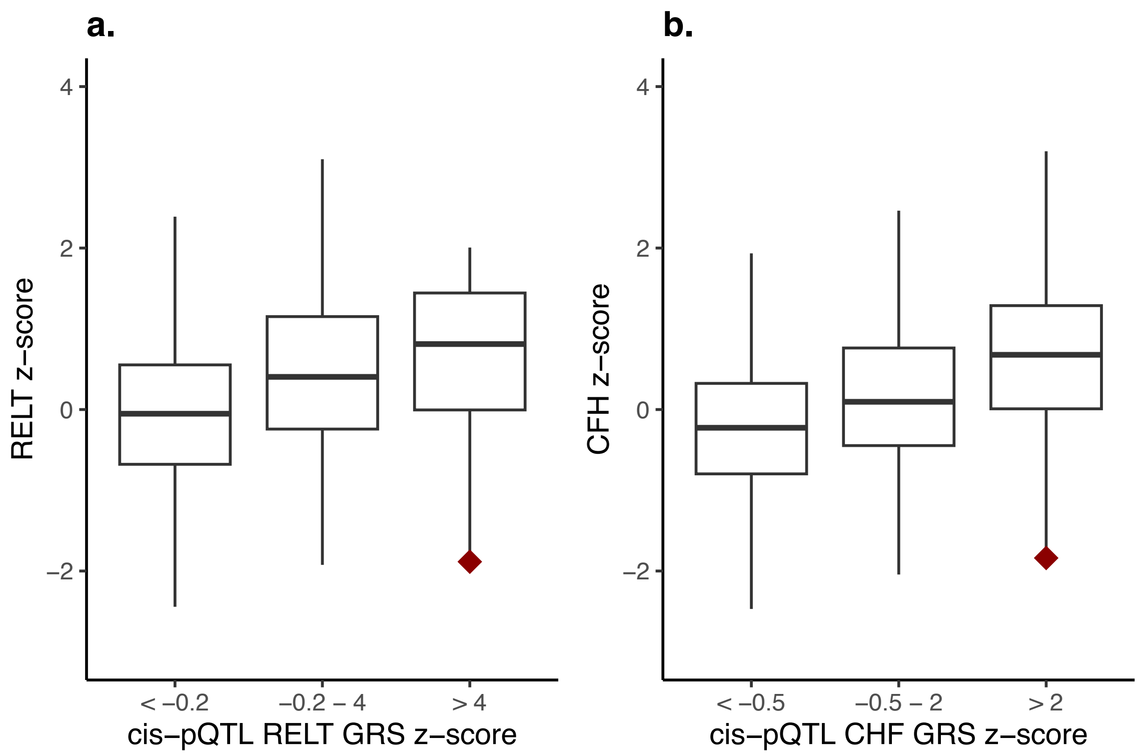


**Supplementary figure 10. Evidence of modifying effects of common protein quantitative trait loci (pQTLs) on cis-protein effects from 'knockout' variants. a,** Distribution of RELT protein abundance across common *RELT* pQTL genetic risk score (GRS) bins. The red diamond shows the RELT protein z-score in an individual carrying a homozygous LoF variant in the *RELT* gene but with a high pQTL GRS (i.e. many protein increasing variants). **b,** Distribution of CHF protein abundance across common *CHF* pQTL genetic risk score (GRS) bins. The red diamond shows the CHF protein z-score in an individual carrying a homozygous LoF variant in the *CHF* gene but with a high pQTL GRS (i.e. many protein increasing variants).

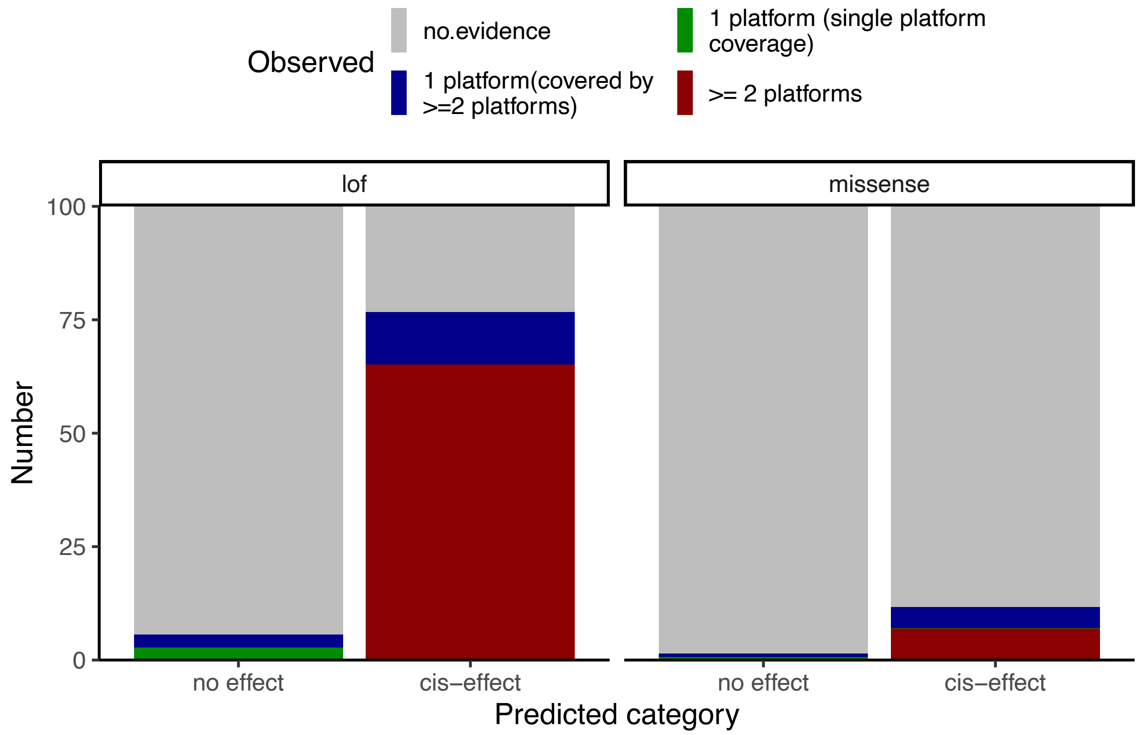


**Supplementary figure 11. Detection rate of 'knockout' variant cis-protein effects for well measured proteins.** We leverage the LoF and missense XGBoost models to predicted the likelihood of observing an effect on *cis*-protein abundance for rare all homozygous or missense variants in genes covered by at least one proteomic platform. We compared the proportion of those variants for which a cis-protein effect was indeed observed between the two predicted variant categories.
